# Adverse childhood experiences and trajectories of psychological distress in adulthood: an analysis of the 1958 British birth cohort

**DOI:** 10.1101/2021.05.19.21257499

**Authors:** Dawid Gondek, Praveetha Patalay, Amanda Sacker, Thierry Gagné, Andrea Danese, Rebecca E Lacey

## Abstract

**Background:** The evidence on the association between adverse childhood experiences (ACEs) and psychological distress in adulthood tends to rely on cross-sectional studies. In this 50-year long longitudinal study, we examined the association between both prospectively and retrospectively reported ACEs and adulthood trajectories of psychological distress between age 23 and 50. We also studied potential effect modifiers of these associations, spanning social and developmental domains of child development.

**Methods:** The sample comprised 8,055 participants of the 1958 National Child Development Study. Psychological distress was measured by the Malaise Inventory at ages 23-50. We used growth curve analysis and multinomial regression.

**Results:** After accounting for covariates, those with more ACEs experienced persistently higher psychological distress between age 23 and 50, with a graded relationship observed. The associations were relatively consistent across prospectively and retrospectively reported ACEs. Those with prospective or retrospective ACE score of 1 compared with 0, had on average between 0.27 and 0.39 higher distress throughout adulthood. In relative terms, the prospective ACE score of 2+ (vs 0) was associated with 3.31 and the retrospective ACE score of 4+ (vs 0) with 5.76 (95% CI 4.24 to 7.82) times higher risk of being in the “high symptoms”, compared with the “low symptoms” trajectory of distress. None of the potential effect modifiers altered the association between ACEs and trajectories of distress.

**Conclusion:** If the associations between ACEs and trajectories of distress are causal, this emphasises the need to act early to prevent psychopathology across the adult life course.

## Introduction

Adverse childhood experiences (ACEs) are broadly defined as potentially traumatic events occurring in childhood and aspects of the child’s environment that can undermine their sense of safety, stability, and bonding (McLaughlin *et al*., 2019). Epidemiological research has shown a dose-response gradient between the number of ACEs (‘ACE score’) and psychological distress (Kessler *et al*., 2010, McLaughlin *et al*., 2010, McLaughlin *et al*., 2012, Selous *et al*., 2020). As ACEs are highly prevalent and largely preventable (McLaughlin *et al*., 2019), acting on them may help to improve population mental health, potentially offsetting rising levels of psychological distress in more recently born cohorts (Gondek *et al*., 2021, Prior *et al*., 2020).

Exposure to ACEs appears to increase vulnerability to psychological distress throughout the entire life course (Ege *et al*., 2015, Selous *et al*., 2020). However, the evidence tends to rely on cross-sectional studies or studies with a short observation period between ACEs and psychological distress, with ACEs typically measured retrospectively and distress assessed only at a single time point (Hughes *et al*., 2017). To our knowledge, only one study included multiple measures of distress across adulthood, finding a persistent association between prospectively measured ACEs and psychological distress between age 23 and 50 in the 1958 British birth cohort (Selous *et al*., 2020). However, the main limitation of the study was that it examined the associations between ACEs and distress at individual ages, without exploring the longitudinal nature of the outcome (Selous *et al*., 2020). This could be an important limitation, as there is evidence for a large variation in trajectories of psychological distress (Prior *et al*., 2020). Moreover, there may be several typical subgroups of distress trajectories in the population, with varying levels of stability over the life course, and with potentially distinct developmental pathways (Colman *et al*., 2007). Hence, in the present study, we used two complementary longitudinal approaches, which not only focus on the average trajectories of distress across age groups but also help to identify subgroups of such trajectories. These subgroups may vary in the magnitude of their association across individuals with a different number of ACEs, hence possibly differing in their aetiology.

In the current study, we included both prospectively and retrospectively reported ACEs, which allows for investigating additional ACEs that were not considered by the previous study using the same data (Selous *et al*., 2020). This includes various forms of abuse, which were reported retrospectively in the 1958 birth cohort, and which tend to be strongly associated with adulthood depression (Mandelli *et al*., 2015). Moreover, prospective and retrospective measures of ACEs tend to identify different groups of individuals, with potentially varying pathways to adult distress (Baldwin *et al*., 2019). Hence, it is important to test the sensitivity of findings to both types of measures.

As the majority of individuals who experience ACEs do not subsequently develop mental health problems, it is important to understand who is either susceptible or resilient to developing high distress after being exposed to ACEs. One approach to shed light on these characteristics is by studying modifiers of the association of ACEs with distress, which either amplify or weaken this relationship (McLaughlin, 2016). There is little evidence on effect modifiers in the context of ACEs and distress in adulthood, with no previous research examining modifiers of the association between ACEs and distress trajectories in adulthood. One study, for instance, showing that low income and female gender may exacerbate the consequences of adversities (Pitkänen *et al*., 2019). In another study, Selous and colleagues found a stronger association between ACEs and distress for men than women at age 50 (Selous *et al*., 2020). Most research typically investigates social, emotional, and neurobiological domains of the child’s development conceptualised as mechanisms (‘mediators’) explaining how ACEs may translate to later psychopathology (Healy *et al*., 2021, Miller *et al*., 2021, Miller *et al*., 2018). In the current study, we identified the potential effect modifiers of ACEs-psychological distress associations according to the criteria – occurring in early-life, spanning multiple domains of early-life development, and having been previously studied in the context of ACEs and mental health, for instance as exposures or mediators (Atkins *et al*., 2020, Barboza Solís *et al*., 2016, Dodgeon *et al*., 2020). They included gender, father’s occupational social class, parental involvement in child’s upbringing, early-life measures of cognitive ability, conscientiousness, internalising problems, externalising problems, and physical health.

In summary, we aimed to expand on previous work by examining the association of ACEs with adult trajectories of psychological distress and by including both prospectively and retrospectively reported ACEs. We examined the association between ACEs and both average trajectories of distress and their subgroups. Finally, we examined various characteristics that may amplify or weaken the association between ACEs and subgroups of distress trajectories.

## Methods

### Study population

The NCDS follows the lives of 17,415 people born in England, Scotland, and Wales in a single week of 1958 (Power and Elliott, 2006). It includes information on physical and educational development, economic circumstances, employment, family life, health behaviours, wellbeing, social participation, and attitudes (Power and Elliott, 2006). The history, design, and features of the NCDS have been described elsewhere (Power and Elliott, 2006). We included only those who completed the retrospective ACEs questionnaire (at age 44/45) (n = 8,146). After excluding those who died, emigrated from Britain, and had no valid measure of psychological distress at any of the data sweeps (by age 50) (n = 91), this resulted in the analytical sample of n = 8,055 (see eFigure 1 for the sample flow diagram). The NCDS was granted ethical approval for each sweep from 2000 by the National Health Service (NHS) Research Ethics Committee and all participants have given informed consent.

**Fig. 1.**
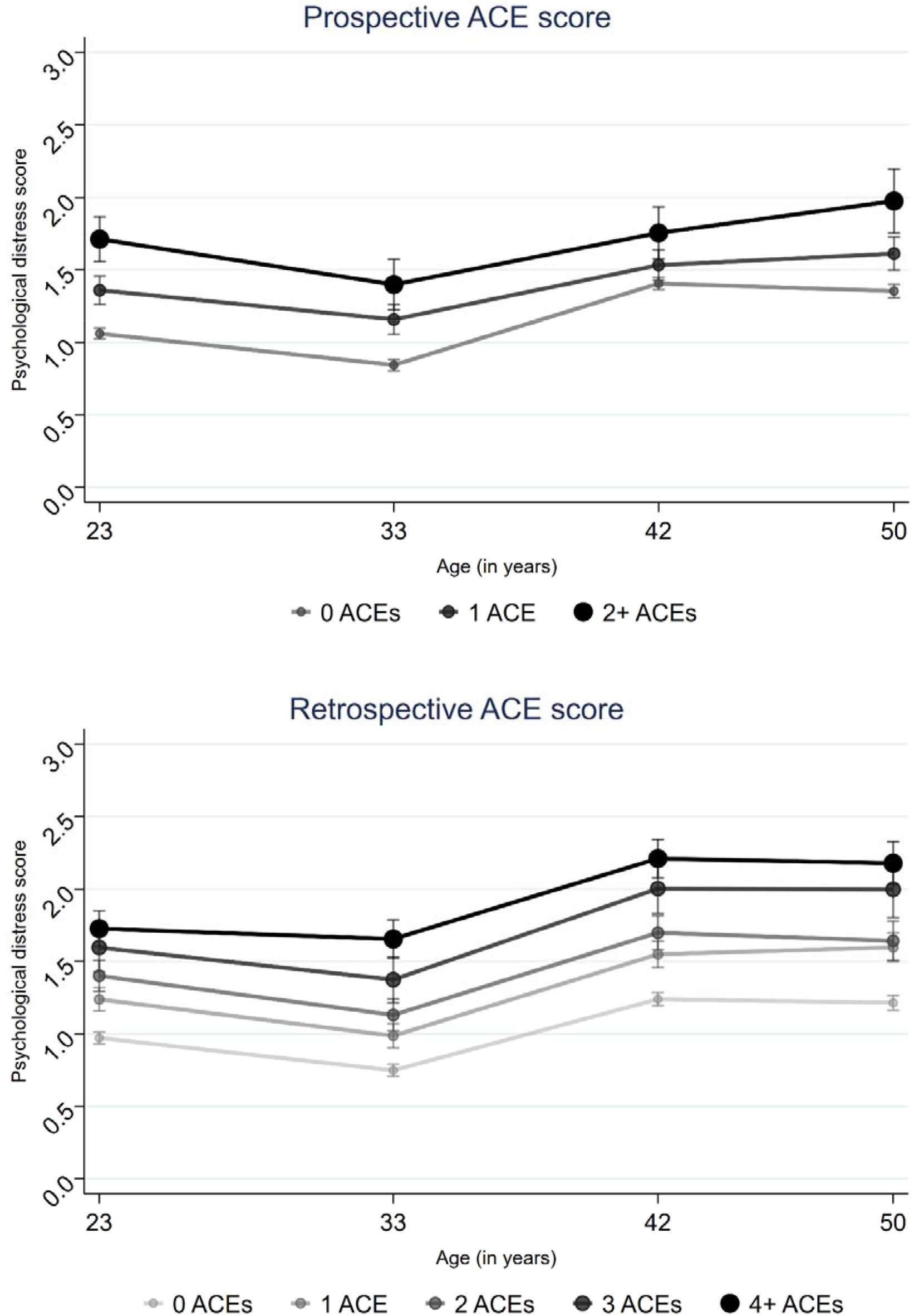
Average trajectories of psychological distress across the ACE score. *Note*. The estimates were adjusted for covariates – father’s occupational social class, maternal education, birthweight, gestational age, maternal age at birth and breastfeeding duration.

### Adverse Childhood Experiences

ACEs were identified based on previous studies using the NCDS, representing traumatic and stressful psychosocial conditions that tend to co-occur, persist over time and are outside of child’s control (Kelly-Irving *et al*., 2013, Lacey *et al*., 2020). ACEs were measured prospectively through reports by parents (usually the mother; at ages 7-16), health visitors or teachers (at ages 7-16) and retrospectively by cohort members (at ages 33 and 44/45). Details on measures of both groups of ACEs can be found in eTable 1. In the current study, ACEs are considered both as an ACE score and individually, the advantages and disadvantages of both approaches were previously discussed in more detail (Lacey and Minnis, 2020, Lacey *et al*., 2020). The prospective ACE score (pACE) was banded as ‘0 pACE’, ‘1 pACE’ and ‘2 + pACEs’ (only 1.3% of cohort members reported 3 or more pACEs). The retrospective ACE score (rACE) was categorised as ‘0 rACE’, ‘1 rACEs’, ‘2 rACEs’, ‘3 rACEs’ and ‘4 + rACEs’, which is consistent with the ACE score literature (Bellis *et al*., 2017, Felitti *et al*., 1998).

**Table 1.**
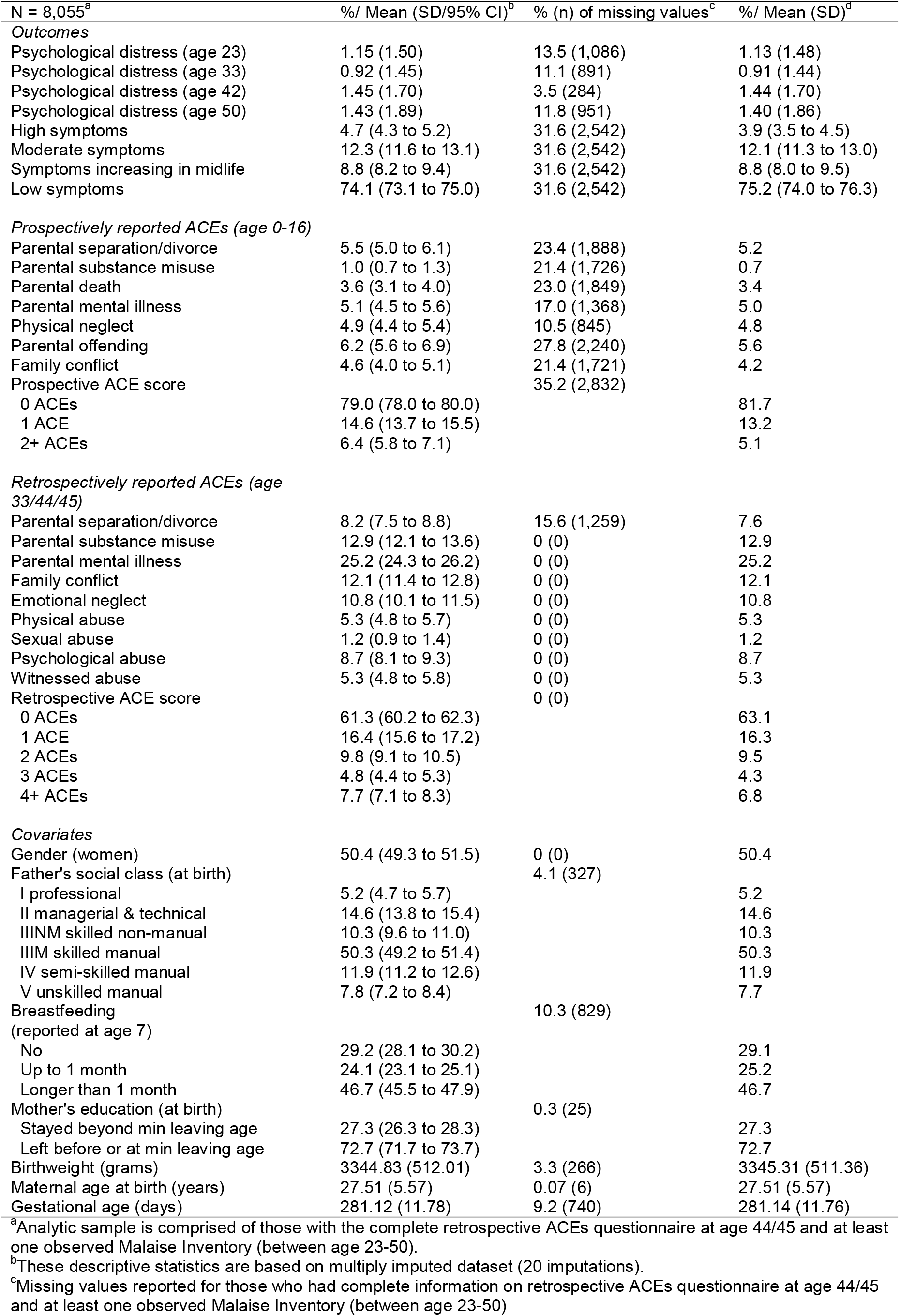
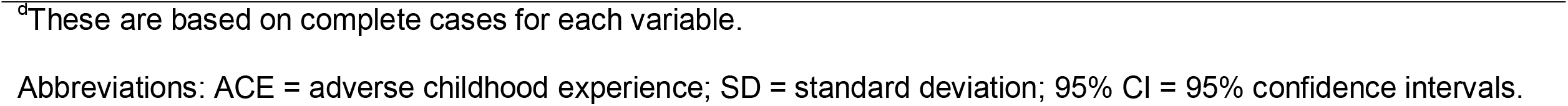
Description of the study sample.

| N = 8,055<sup>a</sup> | %/ Mean (SD/95% CI)<sup>b</sup> | % (n) of missing values<sup>c</sup> | %/ Mean (SD)<sup>d</sup> |
| --- | --- | --- | --- |
| *Outcomes* |  |  |  |
| Psychological distress (age 23) | 1.15 (1.50) | 13.5 (1,086) | 1.13 (1.48) |
| Psychological distress (age 33) | 0.92 (1.45) | 11.1 (891) | 0.91 (1.44) |
| Psychological distress (age 42) | 1.45 (1.70) | 3.5 (284) | 1.44 (1.70) |
| Psychological distress (age 50) | 1.43 (1.89) | 11.8 (951) | 1.40 (1.86) |
| High symptoms | 4.7 (4.3 to 5.2) | 31.6 (2,542) | 3.9 (3.5 to 4.5) |
| Moderate symptoms | 12.3 (11.6 to 13.1) | 31.6 (2,542) | 12.1 (11.3 to 13.0) |
| Symptoms increasing in midlife | 8.8 (8.2 to 9.4) | 31.6 (2,542) | 8.8 (8.0 to 9.5) |
| Low symptoms | 74.1 (73.1 to 75.0) | 31.6 (2,542) | 75.2 (74.0 to 76.3) |
| *Prospectively reported ACEs (age 0-16)* |  |  |  |
| Parental separation/divorce | 5.5 (5.0 to 6.1) | 23.4 (1,888) | 5.2 |
| Parental substance misuse | 1.0 (0.7 to 1.3) | 21.4 (1,726) | 0.7 |
| Parental death | 3.6 (3.1 to 4.0) | 23.0 (1,849) | 3.4 |
| Parental mental illness | 5.1 (4.5 to 5.6) | 17.0 (1,368) | 5.0 |
| Physical neglect | 4.9 (4.4 to 5.4) | 10.5 (845) | 4.8 |
| Parental offending | 6.2 (5.6 to 6.9) | 27.8 (2,240) | 5.6 |
| Family conflict | 4.6 (4.0 to 5.1) | 21.4 (1,721) | 4.2 |
| Prospective ACE score |  | 35.2 (2,832) |  |
| 0 ACEs | 79.0 (78.0 to 80.0) |  | 81.7 |
| 1 ACE | 14.6 (13.7 to 15.5) |  | 13.2 |
| 2+ ACEs | 6.4 (5.8 to 7.1) |  | 5.1 |
| *Retrospectively reported ACEs (age 33/44/45)* |  |  |  |
| Parental separation/divorce | 8.2 (7.5 to 8.8) | 15.6 (1,259) | 7.6 |
| Parental substance misuse | 12.9 (12.1 to 13.6) | 0 (0) | 12.9 |
| Parental mental illness | 25.2 (24.3 to 26.2) | 0 (0) | 25.2 |
| Family conflict | 12.1 (11.4 to 12.8) | 0 (0) | 12.1 |
| Emotional neglect | 10.8 (10.1 to 11.5) | 0 (0) | 10.8 |
| Physical abuse | 5.3 (4.8 to 5.7) | 0 (0) | 5.3 |
| Sexual abuse | 1.2 (0.9 to 1.4) | 0 (0) | 1.2 |
| Psychological abuse | 8.7 (8.1 to 9.3) | 0 (0) | 8.7 |
| Witnessed abuse | 5.3 (4.8 to 5.8) | 0 (0) | 5.3 |
| Retrospective ACE score |  | 0 (0) |  |
| 0 ACEs | 61.3 (60.2 to 62.3) |  | 63.1 |
| 1 ACE | 16.4 (15.6 to 17.2) |  | 16.3 |
| 2 ACEs | 9.8 (9.1 to 10.5) |  | 9.5 |
| 3 ACEs | 4.8 (4.4 to 5.3) |  | 4.3 |
| 4+ ACEs | 7.7 (7.1 to 8.3) |  | 6.8 |
| *Covariates* |  |  |  |
| Gender (women) | 50.4 (49.3 to 51.5) | 0 (0) | 50.4 |
| Father's social class (at birth) |  | 4.1 (327) |  |
| I professional | 5.2 (4.7 to 5.7) |  | 5.2 |
| II managerial & technical | 14.6 (13.8 to 15.4) |  | 14.6 |
| IIINM skilled non-manual | 10.3 (9.6 to 11.0) |  | 10.3 |
| IIIM skilled manual | 50.3 (49.2 to 51.4) |  | 50.3 |
| IV semi-skilled manual | 11.9 (11.2 to 12.6) |  | 11.9 |
| V unskilled manual | 7.8 (7.2 to 8.4) |  | 7.7 |
| Breastfeeding (reported at age 7) |  | 10.3 (829) |  |
| No | 29.2 (28.1 to 30.2) |  | 29.1 |
| Up to 1 month | 24.1 (23.1 to 25.1) |  | 25.2 |
| Longer than 1 month | 46.7 (45.5 to 47.9) |  | 46.7 |
| Mother's education (at birth) |  | 0.3 (25) |  |
| Stayed beyond min leaving age | 27.3 (26.3 to 28.3) |  | 27.3 |
| Left before or at min leaving age | 72.7 (71.7 to 73.7) |  | 72.7 |
| Birthweight (grams) | 3344.83 (512.01) | 3.3 (266) | 3345.31 (511.36) |
| Maternal age at birth (years) | 27.51 (5.57) | 0.07 (6) | 27.51 (5.57) |
| Gestational age (days) | 281.12 (11.78) | 9.2 (740) | 281.14 (11.76) |

<sup>a</sup>Analytic sample is comprised of those with the complete retrospective ACEs questionnaire at age 44/45 and at least one observed Malaise Inventory (between age 23-50).
<sup>b</sup>These descriptive statistics are based on multiply imputed dataset (20 imputations).
<sup>c</sup>Missing values reported for those who had complete information on retrospective ACEs questionnaire at age 44/45 and at least one observed Malaise Inventory (between age 23-50)
<sup>a</sup>These are based on complete cases for each variable.
Abbreviations: ACE = adverse childhood experience; SD = standard deviation; 95% CI = 95% confidence intervals.

### Psychological distress

Psychological distress (symptoms of depression and anxiety) was measured by the Malaise Inventory (Rutter *et al*., 1970). The Malaise Inventory has been found to have good psychometric properties (McGee *et al*., 1986) and has been used in the general population and high-risk groups (Furnham and Cheng, 2015). The scalar invariance of the measure has been found across age and between genders (Ploubidis *et al*., 2019, Ploubidis *et al*., 2017). This implies that the symptoms captured by the items of the Malaise Inventory were interpreted equivalently by participants, regardless of their age, gender or measurement modes used at different ages (Muthén and Asparouhov, 2013). The longer version of the Malaise Inventory, including 24 “yes-no” questions, was used at ages 23, 33, 42. At age 50, 9 of the 24 questions were asked (also in the “yes-no” format). See eTable 2 for items of the questionnaire. To aid comparability, only items on the shorter 9-item version was used in the current study, which correlated highly with the 24-item version (r = 0.89-0.91 at ages 23-42) – resulting in a score from 0 to 9 for each wave.

### Covariates/effect modifiers

The covariates were identified based on previous research on the relationship between ACEs and adult health using NCDS (Chen and Lacey, 2018, Lacey *et al*., 2020, Li *et al*., 2019, Pinto Pereira *et al*., 2017). These included binary gender, father’s occupational social class at birth (‘I professional’, ‘II managerial and technical’, ‘IIINM skilled non-manual’, ‘IIIM skilled manual’, ‘IV semi-skilled manual’ or ‘V unskilled manual’), maternal education (‘mother stayed in education beyond minimum age’ or ‘mother left at/before minimum age’), birthweight (in grams), gestational age (in days), maternal age at birth (in years), and breastfeeding duration (‘none’, ‘up to one month’ or ‘longer than one month’).

The effect modifiers included gender, father’s occupational social class (manual vs non-manual), parental involvement in child’s upbringing (at age 7), cognitive ability (at age 11), conscientiousness, internalising problems, externalising problems, and physical health (all at age 16). eTable 3 includes more details on how the effect modifiers were measured.

### Analytical strategy

We investigated associations between ACEs (as an ACE score and individually) and trajectories of psychological distress in adulthood (age 23-50) using two different approaches to growth curve modelling (Herle *et al*., 2020). The aim of the first approach, multilevel regression analysis, was to examine the association between ACEs and average trajectories of distress across adulthood. Due to a large variation in average trajectories, we subsequently used latent class growth analysis (LCGA) to identify subgroups (or classes) of individuals who shared similar trajectories of psychological distress (Asparouhov and Muthen, 2014). All analyses included men and women combined due to overall consistent patterns in the associations between genders.

### ACEs and average trajectories of psychological distress

The association between ACEs and the average trajectories of distress were studied using multilevel linear regression. This analysis allows for modelling data that are unbalanced in time, including individuals with missing data and accounting for a hierarchical dependency of observations (level 1) within individuals (level 2), with age becoming an observation-level variable (Raudenbush and Bryk, 2002, Suzuki, 2012).

Due to missing information in exposures and covariates, the analysis used data after multiple imputation (MI) by chained equations, generating 20 datasets, which helps to reduce selection bias and maximise power (White *et al*., 2011). More details on the analytical strategy can be found in eAppendix 1 (see for more details on building the models) and eAppendix 2 and eTable 4 (see for missing data strategy). The analysis was conducted in Stata 16 (StataCorp, 2020).

### ACEs and subgroups of trajectories of psychological distress

The association between ACEs and subgroup of distress trajectories was studied in three steps. In the first step, LCGA was conducted using Full Information Maximum Likelihood (FIML) in MPlus Version 8 (Muthen and Muthen, 1997-2017). Models with an increasing number of classes, from two to five, were estimated to identify the optimum solution. Alternative specifications were compared based on model fit indicators – Akaike Information Criterion (AIC), Bayesian Information Criterion (BIC), adjusted Bayesian Information Criterion (adj BIC), (Vuong) Lo–Mendell–Rubin likelihood ratio tests ((V)LMR-LRT) – entropy, the class size (>5%) and interpretability of the classes (Asparouhov and Muthen, 2014, Herle *et al*., 2020). In the second step, due to high entropy (>0.85) – participants were allocated to different classes using the maximum-probability assignment rule according to their posterior probabilities (Asparouhov and Muthen, 2014). In the third step, the classes were then used as a categorical outcome, which was regressed on the number of ACEs using multinomial regression analyses, adjusted for covariates, and based on multiple imputations (20 datasets) (see eAppendix 2 for missing data strategy). We present associations both on a multiplicative scale, as relative risk ratios (RRR), and an additive scale, as predicted probabilities representing average marginal effects. Step three was conducted in Stata 16 (StataCorp, 2020).

#### Effect modification of association between ACEs and trajectories of distress

The potential effect modification of the association between ACEs and classes of distress was examined by including the ACE-score*effect-modifier coefficient in the unadjusted multinomial regressions, and if evidence for effect modification found, adjusted multinomial regressions. This was done separately for each effect modifier. The strength of evidence for effect modification, based on relative risks, was assessed using global Wald tests.

### Sensitivity analyses

Two additional analyses, both based on the adjusted multinomial regression from the step three described above, were conducted to test the robustness of the findings. First, the analysis was rerun to test the sensitivity of the findings to using a less restrictive sample. The sample included those who were not permanent emigrants, were alive by age 50 and had at least two measures of prospectively reported ACEs (n = 13,130; as opposed to n = 8,055 used in the main analysis). Second, the analysis was rerun to account for uncertainty in the class assignment using the posterior probabilities as regression weights (proportional assignment) (Heron *et al*., 2015).

## Results

### Prevalence of ACEs

Having at least one pACE was reported by 21.0%, with 6.4% reporting 2+. Having at least one rACE was reported by 38.7%, with 7.7% reporting 4+. The most prevalent individual ACEs were parental offending (6.2%), parental separation/divorce (5.5%) and parental mental illness (5.1%) among prospective reports, and parental mental illness (25.2%), parental substance misuse (12.9%), and family conflict (12.1%) among retrospective reports.

### ACEs and average trajectories of psychological distress

After accounting for covariates, those with ACEs experienced persistently higher psychological distress between age 23 and 50 (see Figure 1 for trajectories and eTables 5-6 for model estimates), both when ACEs were reported prospectively and retrospectively. The mean difference in psychological distress for 1 pACE vs 0 pACEs ranged from 0.31 (95% CI, 0.20 to 0.42) at age 23 to 0.27 (95% CI, 0.13 to 0.41) at age 50, and for 1 rACE vs 0 rACE from 0.28 (95% CI, 0.17 to 0.36) at age 23 to 0.39 (95% CI, 0.27 to 0.50) at age 50.

Distress increased with the number of ACEs in a graded fashion. For instance, the mean difference in psychological distress for 2+ pACEs vs 1 pACE ranged from 0.35 (95% CI, 0.15 to 0.55) at age 23 to 0.35 (95% CI, 0.09 to 0.60) at age 50, and for 4+ rACE vs 1 rACE from 0.48 (95% CI, 0.32 to 0.64) at age 23 to 0.57 (95% CI, 0.39 to 0.75) at age 50.

There was evidence, in the covariates-adjusted model, for differential slopes across pACE score, but not rACE score (p-value of Wald test for pACE-score*age coefficient = 0.0005 and rACE-score*age coefficient = 0.12). This was due to the increase in distress between age 33 and 42 being greater among those with no ACEs than those with ACEs (e.g., slope at age 33-42: B _0 pACEs_ = 0.062; 95% CI, 0.058 to 0.067 vs B _2+ pACEs_ = 0.041; 95% CI, 0.022 to 0.061), leading to reduced inequality in distress between those with and without pACEs at age 42. The differences increased again until age 50.

Overall, there was substantial variance around trajectories of psychological distress, with the spread of the trajectories widening with age and being shifted towards a higher range of scores among those with 2+ pACE and 4+ rACE scores (see eFigure 4).

The association between ACE scores and trajectories of psychological distress was not driven by any individual ACE, as most ACEs appeared to be strongly associated with distress (see eFigures 2-3).

**Fig. 2.**
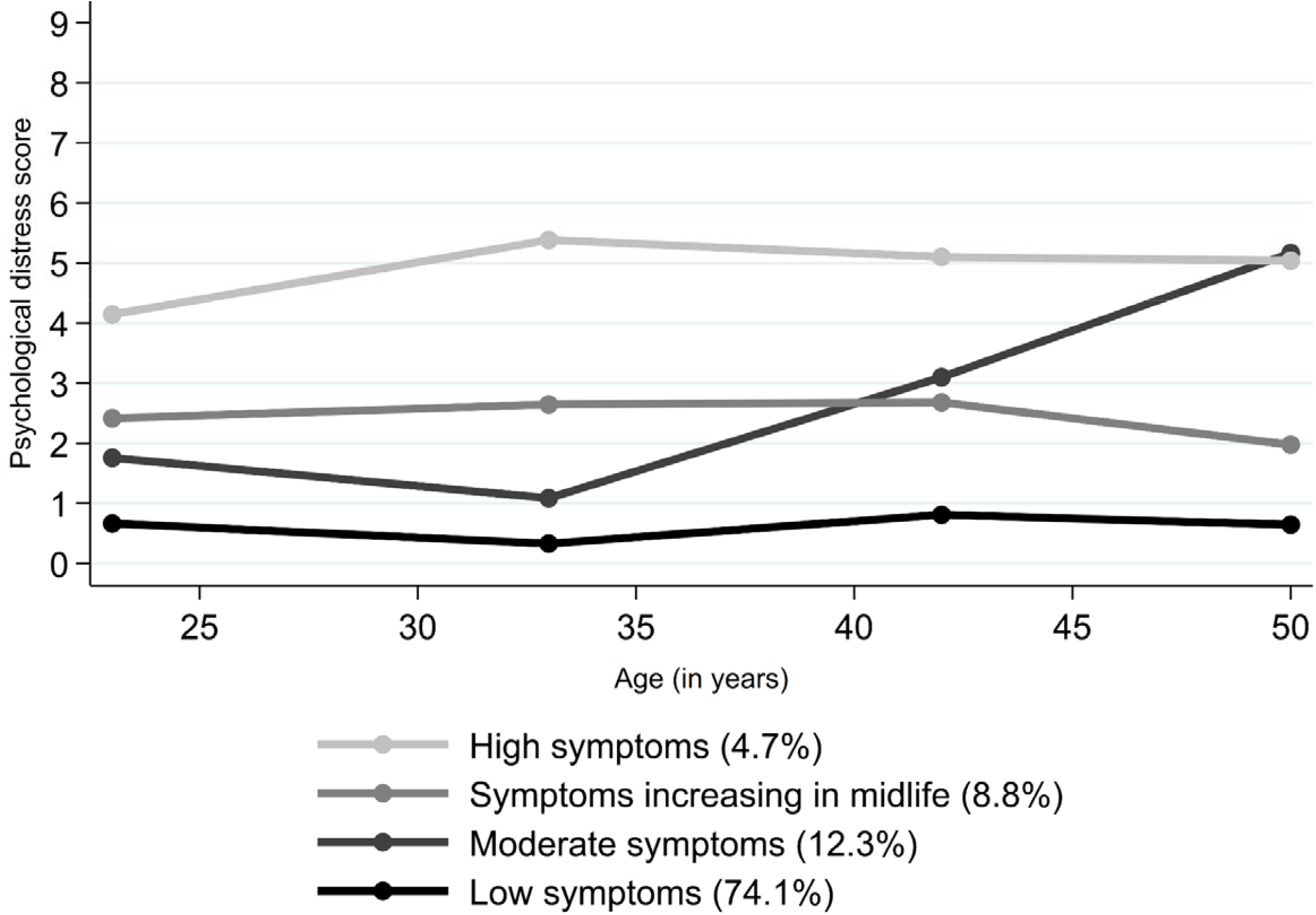
Mean score of psychological distress across age for the four subgroups of distress trajectories.

### ACEs and subgroups of trajectories of psychological distress

The LCGA with four classes was deemed the most suitable solution. The four-class solution showed a better fit than solutions with fewer classes (lower values of AIC, BIC and adj BIC; highly significant LMR-LRT and VLMR-LRT) (see eTable 7), with the 5-class solution not adding any classes of substantial theoretical interest.

The “low symptoms” class was assigned to 74.1% of participants, characterised by a relatively stable mean psychological distress (mean across age = 0.61, standard deviation [SD] = 0.90), 12.3% were assigned to “moderate symptoms” (mean = 2.43, SD=1.36), 4.7% to “high symptoms” (mean = 4.92, SD = 1.96) (see Figure 2). The final class, “symptoms increasing in midlife”, was characterised by a sharp increase in distress between age 34 and 50 (mean = 2.78, SD = 2.15).

As presented in Figure 3, after accounting for covariates, a higher ACE score had a graded association with a lower predicted probability of being in the “low symptoms” and a higher probability of being in one of the three symptomatic classes. For instance, having 2+ pACEs (vs 0) was associated with 14% and 4+ rACEs (vs 0) with 21% lower probability of being in the “low symptoms” class. Having 2+ pACEs (vs 0) was associated with 6% and 4+ rACEs (vs 0) with 7% higher probability of being in the “high symptoms” class.

**Fig. 3.**
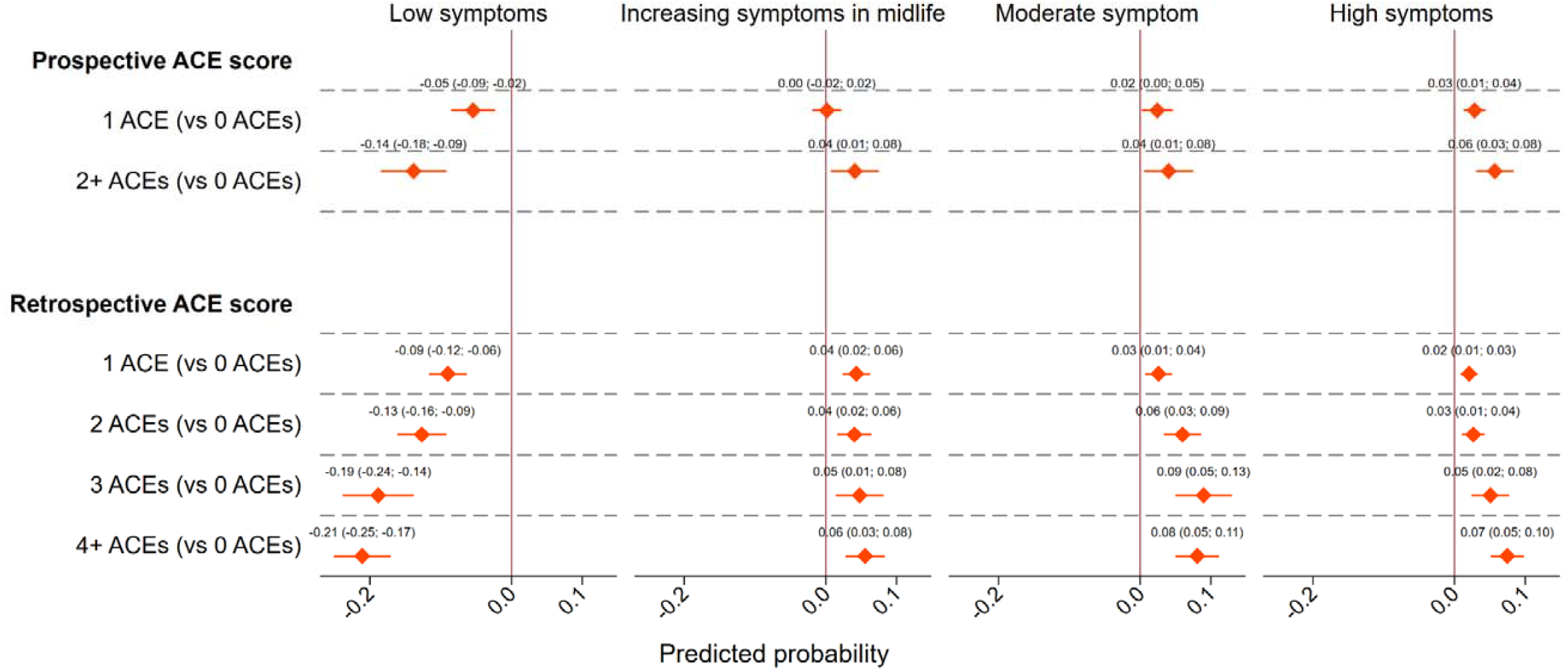
Predicted probabilities of belonging to each subgroup of distress trajectories across ACE score. *Note*. The estimates come from the model adjusted for covariates – gender, father’s occupational social class, maternal education, birthweight, gestational age, maternal age at birth and breastfeeding duration.

In relative terms, 2+ pACEs (vs 0) was associated with 3.31 (95% CI 2.21 to 4.96) and 4+ rACEs (vs 0) with 5.76 (95% CI 4.24 to 7.82) times higher risk of being in the “high symptoms”, compared with the “low symptoms” class (see eTable 8 for associations on a multiplicative scale). These associations were robust to different sample definitions (see eTable 9 for details) and when accounting for uncertainty in the class assignment (see eTable 10 for details).

The family conflict variable had the strongest and most consistent association across all classes, both when reported prospectively and retrospectively (see Figure 4). It was associated with between 13% (reported prospectively) and 18% (reported retrospectively) lower probability of being in the “low symptoms” class and between 11% (reported prospectively) and 14% (reported retrospectively) higher probability of being in the “moderate symptoms” or “high symptoms” classes. Parental death or separation had the weakest association with all classes. The associations were relatively consistent in magnitude among retrospectively reported individual ACEs, with all forms of abuse and neglect having a particularly strong link with the “low symptoms” and “moderate/high symptoms” classes (see Figure 4).

**Fig. 4.**
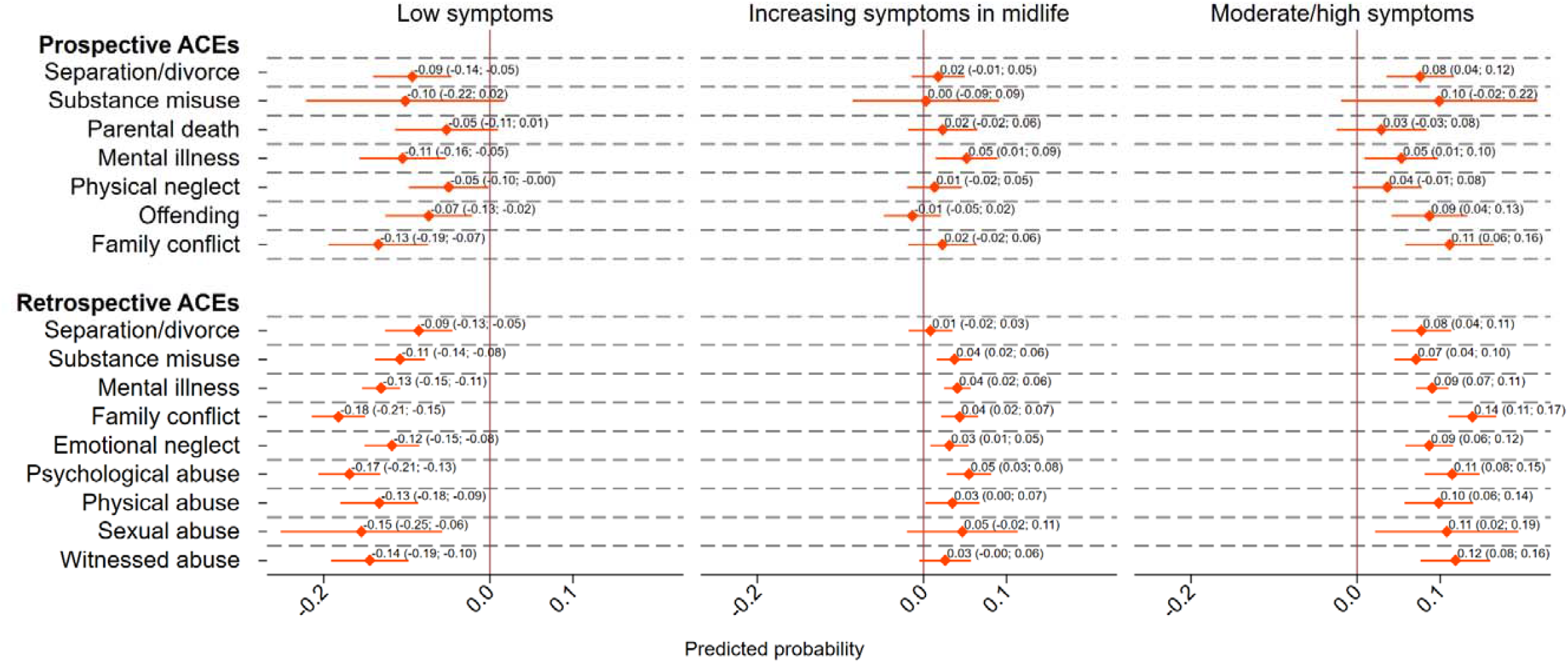
Differences in predicted probabilities of belonging to each subgroup of distress trajectories across individual ACEs. *Note*. The estimates come from the model adjusted for covariates – gender, father’s occupational social class, maternal education, birthweight, gestational age, maternal age at birth and breastfeeding duration. “Moderate symptoms” and “high symptoms” subgroups were combined due to similar patterns of associations across both groups and to increase the statistical power of the analysis.

#### Effect modification of association between ACEs and trajectories of distress

All studied effect modifiers, including father’s occupational social class (manual vs non-manual), gender, parental involvement (at age 7), cognitive ability (at age 11), conscientiousness, internalising problems, externalising problems, physical health problem (all at age 16), were strongly associated with subgroups of distress trajectories (see eTable 11). However, we found no evidence for effect modification of the association between the ACE score, reported either prospectively or retrospectively, and subgroups of distress trajectories. In unadjusted models, the p-values of Wald tests ranged between 0.08 (for rACE score*cognitive ability) and 0.94 (for pACE score*social class).

## Discussion

### Summary of the findings in the context of previous research

Building on the body of work that has examined the age-specific associations between ACEs and psychological distress in NCDS, we investigated the link between ACEs and adulthood trajectories of psychological distress between ages 23 and 50 (Selous *et al*., 2020). In line with previous literature (Kessler *et al*., 2010, McLaughlin *et al*., 2010, McLaughlin *et al*., 2012, Selous *et al*., 2020), after accounting for covariates, those with more ACEs experienced persistently higher psychological distress, with a graded relationship.

Consistent with previous studies in the United Kingdom (Bell, 2014, Colman *et al*., 2007, Prior *et al*., 2020), we found large variation in individual trajectories of psychological distress throughout adulthood, hence we identified subgroups of these trajectories. To our knowledge, such subgroups have not been studied previously in the context of adversity. Across the identified subgroups, individuals with increasingly higher ACE scores were proportionally less likely to be in the “low symptoms” class, and more likely to be in the three other symptomatic subgroups. The risk of being in the “high symptoms” compared to the “low symptoms” subgroup was nearly six times higher among those with four or more (versus none) retrospectively reported ACEs.

None of the potential effect modifiers we examined (gender, father’s occupational social class, parental involvement, cognitive ability, conscientiousness, internalising problems, externalising problems, and physical health problems) altered the association between ACEs and trajectories of distress. Previously, these social and developmental domains have been found to partially explain the association between ACEs and psychopathology (Healy *et al*., 2021, Miller *et al*., 2021, Miller *et al*., 2018). However, they do not appear to amplify or weaken the relationship between ACEs and adult trajectories of psychological distress.

All studied individual ACEs were associated with higher levels of psychological distress, with family conflict having the strongest and most consistent relationship with all classes, both when reported prospectively and retrospectively. On the other hand, parental death or separation had the weakest association with classes of distress. The association between family conflict and distress was not studied in this cohort before. However, it was found to have a strong association with inflammatory markers in midlife (Lacey *et al*., 2020). Various forms of abuse and neglect, here reported retrospectively, were previously found to be strongly associated with adulthood depression (Mandelli *et al*., 2015), recurrent and persistent depressive episodes and lack of response or remission during treatment for depression (Nanni *et al*., 2012).

### Interpretation and implications of the findings

Several potential mechanisms may explain the link between ACEs and psychological distress. Our findings emphasise the stability of the relationship between ACEs and psychological distress during adulthood. This could be due to ACEs leading to physiological dysregulation of the adrenal axis or immune system associated with chronic stress (McLaughlin *et al*., 2019). This mechanism may play a particularly important role for experiences involving harm or threat of harm, such as family conflict, which are more likely to activate the hypothalamic-pituitary-adrenal axis (McLaughlin *et al*., 2019). The importance of preventing family conflict has been recently recognised by the Early Intervention Foundation, which outlines recommendations on how the population needs concerning parental conflict can be assessed and targeted at a local level (Ghiara *et al*., 2021).

Although ACEs are strongly associated with the risk of subsequent distress, most individuals with a high ACE score maintain low symptoms throughout their adulthood. Understanding what differentiates those who experienced ACEs and subsequently suffered high distress from those who remained resilient would help to identify individuals at elevated risk and learn about characteristics that could be nurtured for secondary prevention against distress. We studied various aspects of development that may amplify or weaken the association between ACEs and subgroups of distress trajectories, finding no such effect.

There are several potential explanations for this somewhat unexpected finding. First, one might speculate that the studied characteristics could have a modifying effect on distress at specific ages, but not trajectories of distress throughout adulthood due to interactive effects fading away with age. Hence, finding an effect would seem most likely for distress at age 23, as it is the closest in time to measurement of effect modifiers that were collected by age 16. We have tested this post-hoc hypothesis, and we found evidence for the interactive protective effect of cognitive ability for the association between retrospectively reported ACEs and distress at age 23 and 33, but not at later ages (results not shown). However, these results need to be interpreted with caution as multiple testing for effect modification at each age increases the chances of finding false-positive effects. Another plausible explanation for the lack of evidence on effect modification is insufficient power resulting from categorising both exposures and the outcome. However, when we used the ACE score as a continuous exposure, to test this speculative hypothesis, we obtained similar results. Future research might consider a range of other potential effect modifiers. For instance, social support was previously found to moderate the influence of parental psychopathology on child’s depressive symptoms (Klasen *et al*., 2015). Also, effect modification of factors more proximal to adult measures of distress could be tested, for instance, related to relationships in young adulthood, education and employment (Hatch, 2005).

Prospectively and retrospectively reported ACEs tend to correlate poorly hence potentially identifying different groups of individuals (Baldwin *et al*., 2019, Newbury *et al*., 2018). One key difference in findings on both types of measures was reduced association between prospectively, but not retrospectively, reported ACEs and distress at age 42. Individuals with multiple retrospectively reported ACEs peaked in their distress at age 42, whereas those with prospectively reported ACEs experienced a further rise in distress until age 50. As high distress correlates with a greater propensity to report ACEs retrospectively (Baldwin *et al*., 2019, Colman *et al*., 2016, Reuben *et al*., 2016), high distress at age 42 likely might have contributed to overreporting ACEs retrospectively at age 44/45. Unfortunately, direct comparison of consistency between the prospective and retrospective reports was not possible, due to little overlap in individual ACEs measured in both ways. For ACEs that were measured both prospectively and retrospectively, such as parental separation or divorce and mental illness, we found relatively consistent associations. The main advantage of using retrospective reports was that it allowed for including additional ACEs, not previously considered in the context of mental health using the 1958 birth cohort, such as different forms of abuse.

### Strengths and limitations

The main strength of our study was that it explored a wide range of ACEs, considered both as an ACE score and individually while accounting for the longitudinal nature of psychological distress. This is important due to non-linear and highly diverse trajectories of distress in adulthood. A potential limitation is that the measure of distress – the Malaise Inventory – does not identify clinical diagnoses. However, it has good predictive validity of the interview-assessed diagnosis of depression and use of mental health services (Clark *et al*., 2010, Rodgers *et al*., 1999).

Our sample is broadly representative of British men and women born around 1958. However, missing data due to non-response and attrition may introduce bias. To mitigate this limitation, we used FIML and multiple imputation. These approaches assume that the data are missing at random, implying that systematic differences between the missing and the observed values can be explained by observed data (Collins *et al*., 2001). This assumption is largely untestable, however, it can be maximised by including auxiliary variables in the imputation model (Sterne *et al*., 2009). These are widely available in the 1958 British birth cohort and were used in FIML and multiple imputation in this study, prediction of missing values with greater precision and accuracy (Mostafa *et al*., 2021, Mostafa and Wiggins, 2015). Moreover, the main findings were consistent when using a less restrictive sample, which is likely to be more representative of the general population but also suffers from a greater proportion of missing information.

### Conclusions

Adverse childhood experiences are associated, in a graded fashion, with trajectories of increasing and sustained distress from young to middle adulthood. If associations are causal, this emphasises the need to act early to prevent psychopathology across the adult life course.

## Data Availability

Cohort data comply with ESRC data sharing policies, readers can access data via the UK Data Archive (www.data-archive.ac.uk), through a formal request.

## Author Contributions

Dawid Gondek had full access to all of the data in the study and takes responsibility for the integrity of the data and the accuracy of the data analysis.

Study concept and design: Dawid Gondek, Rebecca Lacey, Praveetha Patalay.

Acquisition, analysis, or interpretation of data: Dawid Gondek.

Drafting of the manuscript: Dawid Gondek.

Critical revision of the manuscript for important intellectual content: Dawid Gondek, Praveetha Patalay, Andrea Danese, Thierry Gagné, Amanda Sacker, Rebecca Lacey.

Statistical analysis: Dawid Gondek.

Administrative, technical, or material support: Dawid Gondek, Praveetha Patalay, Andrea Danese, Thierry Gagné, Amanda Sacker, Rebecca Lacey.

Study supervision: Dawid Gondek, Praveetha Patalay, Rebecca Lacey.

## Acknowledgments

We would like to thank all the cohort members of the 1958 National Child Development Study, who generously gave their time to participate in this project and without whom this research resource would not now be available.

## References

Asparouhov, T. & Muthen, B. (2014). Auxiliary Variables in Mixture Modeling: Three-Step Approaches Using Mplus. Structural Equation Modeling 21, 329–41.

Atkins, R., Turner, A. J., Chandola, T. & Sutton, M. (2020). Going beyond the mean in examining relationships of adolescent non-cognitive skills with health-related quality of life and biomarkers in later-life. Economics & Human Biology 39, 100923.

Baldwin, J. R., Reuben, A., Newbury, J. B. & Danese, A. (2019). Agreement between prospective and retrospective measures of childhood maltreatment. JAMA Psychiatry.

Barboza Solís, C., Fantin, R., Castagné, R., Lang, T., Delpierre, C. & Kelly-Irving, M. (2016). Mediating pathways between parental socio-economic position and allostatic load in mid-life: Findings from the 1958 British birth cohort. Social Science & Medicine 165, 19–27.

Bell, A. (2014). Life-course and cohort trajectories of mental health in the UK, 1991-2008--a multilevel age-period-cohort analysis. Soc Sci Med 120, 21–30.

Bellis, M., Hughes, K., Hardcastle, K., Ashton, K., Ford, K., Quigg, Z. & Davies, A. (2017). The impact of adverse childhood experiences on health service use across the life course using a retrospective cohort study. Journal of Health Services Research & Policy 22, 168–17.

Chen, M. & Lacey, R. E. (2018). Adverse childhood experiences and adult inflammation: Findings from the 1958 British birth cohort. Brain Behav Immun 69, 582–590.

Clark, C., Caldwell, T., Power, C. & Stansfeld, S. (2010). Does the influence of childhood adversity on psychopathology persist across the lifecourse? A 45-year prospective epidemiologic study. Annals of Epidemiology 20, 385–394.

Collins, L. M., Schafer, J. L. & Kam, C. M. (2001). A comparison of inclusive and restrictive strategies in modern missing data procedures. Psychological Methods 6, 330–351.

Colman, I., Kingsbury, M., Garad, Y., Zeng, Y., Naicker, K., Patten, S., Jones, P. B., Wild, T. C. & Thompson, A. H. (2016). Consistency in adult reporting of adverse childhood experiences. Psychological Medicine 46, 543–549.

Colman, I., Ploubidis, G. B., Wadsworth, M. E., Jones, P. B. & Croudace, T. J. (2007). A longitudinal typology of symptoms of depression and anxiety over the life course. Biol Psychiatry 62, 1265–71.

Dodgeon, B., Patalay, P., Ploubidis, G. B. & Wiggins, R. D. (2020). Exploring the role of early-life circumstances, abilities and achievements on well-being at age 50 years: evidence from the 1958 British birth cohort study. BMJ Open 10, e031416.

Ege, M. A., Messias, E., Thapa, P. B. & Krain, L. P. (2015). Adverse childhood experiences and geriatric depression: results from the 2010 BRFSS. Am J Geriatr Psychiatry 23, 110–4.

Felitti, V., Anda, R. F., Nordenberg, D., Williamson, D. F., Spitz, A. M., Edwards, V., Koss, M. P. & Marks, J. S. (1998). Relationship of childhood abuse and household dysfunction to many of the leading causes of death in adults. American Journal of Preventive Medicine 14, 245–258.

Furnham, A. & Cheng, H. (2015). The stability and change of malaise scores over 27 years: Findings from a nationally representative sample. Personality and Individual Differences 79, 30–34.

Ghiara, V., Lewing, B., Chapman, S. & Bethel, C. (2021). Conducting a needs assessment on parental conflict. A step-by-step guide. Early Intervention Foundation.

Gondek, D., Bann, D., Patalay, P., Goodman, A., Richards, M. & Ploubidis, G. B. (2021). Psychological distress from adolescence to early old age: Evidence from the 1946, 1958 and 1970 British birth cohorts. Psychological Medicine.

Hatch, S. L. (2005). Conceptualizing and identifying cumulative adversity and protective resources: implications for understanding health inequalities. J Gerontol B Psychol Sci Soc Sci 60 Spec No 2, 130–4.

Healy, C., Eaton, A., Cotter, I., Carter, E., Dhondt, N. & Cannon, M. (2021). Mediators of the longitudinal relationship between childhood adversity and late adolescent psychopathology. Psychological Medicine, 1–9.

Herle, M., Micali, N., Abdulkadir, M., Loos, R., Bryant-Waugh, R., Hubel, C., Bulik, C. M. & De Stavola, B. L. (2020). Identifying typical trajectories in longitudinal data: modelling strategies and interpretations. Eur J Epidemiol 35, 205–222.

Heron, J., Croudace, T. J., Barker, E. D. & Tilling, K. (2015). A comparison of approaches for assessing covariate effects in latent class analysis. Longitudinal and Life Course Studies 6, 420–434.

Hughes, K., Bellis, M. A., Hardcastle, K. A., Sethi, D., Butchart, A., Mikton, C., Jones, L. & Dunne, M. P. (2017). The effect of multiple adverse childhood experiences on health: a systematic review and meta-analysis. Lancet Public Health 2, e356–e366.

Kelly-Irving, M., Lepage, B., Dedieu, D., Lacey, R., Cable, N., Bartley, M., Blane, D., Grosclaude, P., Lang, T. & Delpierre, C. (2013). Childhood adversity as a risk for cancer: findings from the 1958 British birth cohort study. BMC Public Health 13, 767.

Kessler, R. C., McLaughlin, K. A., Green, J. G., Gruber, M. J., Sampson, N. A., Zaslavsky, A. M., Aguilar-Gaxiola, S., Alhamzawi, A. O., Alonso, J., Angermeyer, M., Benjet, C., Bromet, E., Chatterji, S., de Girolamo, G., Demyttenaere, K., Fayyad, J., Florescu, S., Gal, G., Gureje, O., Haro, J. M., Hu, C. Y., Karam, E. G., Kawakami, N., Lee, S., Lepine, J. P., Ormel, J., Posada-Villa, J., Sagar, R., Tsang, A., Ustun, T. B., Vassilev, S., Viana, M. C. & Williams, D. R. (2010). Childhood adversities and adult psychopathology in the WHO World Mental Health Surveys. Br J Psychiatry 197, 378–85.

Klasen, F., Otto, C., Kriston, L., Patalay, P., Schlack, R., Ravens-Sieberer, U. & group, B. s. (2015). Risk and protective factors for the development of depressive symptoms in children and adolescents: results of the longitudinal BELLA study. Eur Child Adolesc Psychiatry 24, 695–703.

Lacey, R. & Minnis, H. (2020). Practitioner Review: Twenty years of research with Adverse Childhood Experience (ACE) scores: advantages, disadvantages and applications to practice. Journal of Child Psychology and Psychiatry 61, 116–130.

Lacey, R. E., Pinto Pereira, S. M., Li, L. & Danese, A. (2020). Adverse childhood experiences and adult inflammation: Single adversity, cumulative risk and latent class approaches. Brain Behav Immun 87, 820–830.

Li, L., Pinto Pereira, S. M. & Power, C. (2019). Childhood maltreatment and biomarkers for cardiometabolic disease in mid-adulthood in a prospective British birth cohort: associations and potential explanations. BMJ Open 9, e024079.

Mandelli, L., Petrelli, C. & Serretti, A. (2015). The role of specific early trauma in adult depression: A meta-analysis of Published literature. European Psychiatry 30, 665–680.

McGee, R., Williams, S. & Silva, P. A. (1986). An evaluation of the Malaise inventory. Journal of Psychosomatic Research 30, 147–152.

McLaughlin, K. A. (2016). Future Directions in Childhood Adversity and Youth Psychopathology. J Clin Child Adolesc Psychol 45, 361–82.

McLaughlin, K. A., Green, J. G., Gruber, M. J., Sampson, N. A., Zaslavsky, A. M. & Kessler, R. C. (2010). Childhood adversities and adult psychopathology in the National Comorbidity Survey Replication (NCS-R) III: associations with functional impairment related to DSM-IV disorders. Psychol Med 40, 847–59.

McLaughlin, K. A., Greif Green, J., Gruber, M. J., Sampson, N. A., Zaslavsky, A. M. & Kessler, R. C. (2012). Childhood adversities and first onset of psychiatric disorders in a national sample of US adolescents. Arch Gen Psychiatry 69, 1151–60.

McLaughlin, K. A., Weissman, D. & Bitran, D. (2019). Childhood Adversity and Neural Development: A Systematic Review. Annu Rev Dev Psychol 1, 277–312.

Miller, A. B., Machlin, L., McLaughlin, K. A. & Sheridan, M. A. (2021). Deprivation and psychopathology in the Fragile Families Study: A 15-year longitudinal investigation. J Child Psychol Psychiatry 62, 382–391.

Miller, A. B., Sheridan, M. A., Hanson, J. L., McLaughlin, K. A., Bates, J. E., Lansford, J. E., Pettit, G. S. & Dodge, K. A. (2018). Dimensions of deprivation and threat, psychopathology, and potential mediators: A multi-year longitudinal analysis. J Abnorm Psychol 127, 160–170.

Mostafa, T., Narayanan, M., Pongiglione, B., Dodgeon, B., Goodman, A., Silverwood, R. & Ploubidis, G. (2021). Missing at random assumption made more plausible: evidence from the 1958 British birth cohort. Journal of Clinical Epidemiology.

Mostafa, T. & Wiggins, R. D. (2015). The impact of attrition and non-response in birth cohort studies: a need to incorporate missingness strategies. Longitudinal and Life Course Studies 6, 131–146.

Muthén, B. & Asparouhov, T. (2013). New methods for the study of measurement invariance with many groups. Mplus (https://www.statmodel.com/download/PolAn.pdf).

Muthen, L. K. & Muthen, B. (1997-2017). Mplus User’s Guide. Eighth Edition. Muthén & Muthén: Los Angeles, CA.

Nanni, V., Uher, R. & Danese, A. (2012). Childhood maltreatment predicts unfavorable course of illness and treatment outcome in depression: a meta-analysis. Am J Psychiatry 169, 141–51.

Newbury, J. B., Arseneault, L., Moffitt, T. E., Caspi, A., Danese, A., Baldwin, J. R. & Fisher, H. L. (2018). Measuring childhood maltreatment to predict early-adult psychopathology: Comparison of prospective informant-reports and retrospective self-reports. J Psychiatr Res 96, 57–64.

Pinto Pereira, S. M., Li, L. & Power, C. (2017). Child Maltreatment and Adult Living Standards at 50 Years. Pediatrics 139, e20161595.

Pitkänen, J., Remes, H., Aaltonen, M. & Martikainen, P. (2019). Experience of maternal and paternal adversities in childhood as determinants of self-harm in adolescence and young adulthood. Journal of Epidemioly and Community Health 73, 1040–1046.

Ploubidis, G., McElroy, E. & Moreira, H. (2019). A longitudinal examination of the measurement equivalence of mental health assessments in two British birth cohorts. Longitudinal and Life Course Studies 10, 471–489.

Ploubidis, G. B., Sullivan, A., Brown, M. & Goodman, A. (2017). Psychological distress in mid-life: evidence from the 1958 and 1970 British birth cohorts. Psychological Medicine 47, 291–303.

Power, C. & Elliott, J. (2006). Cohort profile: 1958 British birth cohort (National Child Development Study). Int J Epidemiol 35, 34–41.

Prior, L., Jones, K. & Manley, D. (2020). Ageing and cohort trajectories in mental ill-health: An exploration using multilevel models. PLoS One 15, e0235594.

Raudenbush, S. W. & Bryk, A. S. (2002). Hierarchical linear models: Applications and data analysis methods (2nd ed.). Sage Publications: Thousand Oaks.

Reuben, A., Moffitt, T. E., Caspi, A., Belsky, D. W., Harrington, H., Schroeder, F., Hogan, S., Ramrakha, S., Poulton, R. & Danese, A. (2016). Lest we forget: comparing retrospective and prospective assessments of adverse childhood experiences in the prediction of adult health. Journal of Child Psychology and Psychiatry 57, 1103–1112.

Rodgers, B., Pickles, A., Power, C., Collishaw, S. & Maughan, B. (1999). Validity of the Malaise Inventory in general population samples. Social Psychiatry and Psychiatric Epidemiology 34, 333–41.

Rutter, M., Tizard, J. & Whitmore, K. (1970). Education, Health and Behaviour. Longmans: London.

Selous, C., Kelly-Irving, M., Maughan, B., Eyre, O., Rice, F. & Collishaw, S. (2020). Adverse childhood experiences and adult mood problems: evidence from a five-decade prospective birth cohort. Psychol Med 50, 2444–2451.

StataCorp (2020). Stata Statistical Software: Release 16. StataCorp LLC: College Station, TX.

Sterne, J. A., White, I. R., Carlin, J. B., Spratt, M., Royston, P., Kenward, M. G., Wood, A. M. &

Carpenter, J. R. (2009). Multiple imputation for missing data in epidemiological and clinical research: potential and pitfalls. BMJ 338, b2393.

Suzuki, E. (2012). Time changes, so do people. Social Science & Medicine 75, 452–456.

White, I. R., Royston, P. & Wood, A. M. (2011). Multiple imputation using chained equations: issues and guidance for practice. Stat Med. 30, 377–99.

