## Supplemental material for "Adverse childhood experiences and trajectories of psychological distress in adulthood: an analysis of the 1958 British birth cohort"

**Supplementary Material**

Contents

eFigure 1. Sample flow diagram	2

eFigure 2. Trajectories of psychological distress across participants with individual prospectively reported ACEs.	12

eFigure 3. Trajectories of psychological distress across participants with individual retrospectively reported ACEs.	13

eFigure 4. Span of individual age trajectories of psychological distress.	14

**Did not complete retrospective ACE questionnaire (at age 44/45):** n = 9,269

**Original sample**

**(born in Great Britain):**

n = 17,415

**Died or emigrated by the last data sweep (age 50):** n = 91

n = 8,146

n = 8,055

**Did not have any outcome measure (by age 50):** n = 0

**Analytical sample:**

n = 8,055

### eFigure 1. Sample flow diagram

| eTable 1. Measures of prospectively and retrospectively reported adverse childhood experiences (ACEs). | | | |
| --- | --- | --- | --- |
| **Adversity** | **Age collected** | **Informant** | **Description** |
| **Prospectively reported ACEs** | | | |
| Parental separation/divorce | 7 | Health visitor | Divorce or separation listed as difficulty of the family |
|  | 11, 16 | Parent | Relationship to child of person acting as child's parents & reason for change |
| Parental substance misuse | 7 | Health visitor | Alcoholism listed as difficulty of the family |
| Family conflict | 7 | Health visitor | Domestic tension listed as difficulty of the family |
| Death of parent | 7 | Health visitor | Death of mother listed as difficulty of the family |
|  | 7 | Health visitor | Death of child's father listed as difficulty of the family |
|  | 11, 16 | Parent | Relationship to child of person acting as child's parents & reason for change |
| Parental mental health problems | 7 | Health visitor | Mental illness or neurosis listed as difficulty of the family |
|  | 7 | Health visitor | Family uses services of a psychiatric social worker |
|  | 7, 11, 16 | Parent | Mother or father has a chronic mental illness |
| Physical neglect | 7, 11 | Teacher | Child appears scruffy/dirty/underfed |
| Parental offending | 7 | Health visitor | Family in contact with probation services |
|  | 11 | Parent | Family in contact with probation services or family member in prison |
|  | 16 | Parent | Family member in contact with probation services |
| **Retrospectively reported ACEs** | | | |
| Parental separation/divorce | 33 | Cohort member | Parents ever permanently separated or divorced & how old when happened |
| Parental substance misuse | 44/45 | Cohort member | Mother had trouble with drinking or other drug use |
|  | 44/45 | Cohort member | Father had trouble with drinking or other drug use |
| Family conflict | 44/45 | Cohort member | There was much conflict and tension in the household whilst I was growing up |
| Witnessing abuse | 44/45 | Cohort member | I witnessed physical or sexual abuse of others in my family |
| Parental mental health problems | 44/45 | Cohort member | Mother or father suffered from nervous or emotional trouble or depression |
| Sexual abuse | 44/45 | Cohort member | I was sexually abused by a parent |
| Physical abuse | 44/45 | Cohort member | I was physically abused by a parent - punched, kicked or hit or beaten with an objective, or needed medical treatment |
| Psychological abuse | 44/45 | Cohort member | I was verbally abused by a parent |
|  | 44/45 | Cohort member | I suffered humiliation, ridicule, bullying or mental cruelty from a parent |
| Emotional neglect | 44/45 | Cohort member | My mother was unaffectionate |
|  | 44/45 | Cohort member | My father was unaffectionate |

### eTable 2. Items of the Malaise Inventory.

How are you feeling generally…

1. Do you often have backache?

**2. Do you feel tired most of the time?**

**3. Do you often feel miserable or depressed?**

4. Do you often have bad headaches?

**5. Do you often get worried about things?**

6. Do you usually have great difficulty in falling or staying asleep?

7. Do you usually wake unnecessarily early in the morning?

8. Do you wear yourself out worrying about your health?

**9. Do you often get in a violent rage?**

10. Do people often annoy and irritate you?

11. Have you at times had twitching of the face, head or shoulders?

**12. Do you often suddenly become scared for no good reason?**

13. Are you scared to be alone when there are no friends near you?

**14. Are you easily upset or irritated?**

15. Are you frightened of going out alone or of meeting people?

**16. Are you constantly keyed up and jittery?**

17. Do you suffer from indigestion?

18. Do you suffer from an upset stomach?

19. Is your appetite poor?

**20. Does every little thing get on your nerves and wear you out?**

**21. Does your heart often race like mad?**

22. Do you often have bad pains in your eyes?

23. Are you troubled with rheumatism or fibrositis?

24. Have you ever had a nervous breakdown?

*Note.* In bold – nine items used across all ages (23-50).

| eTable 3. Measures of potential effect modifiers. | | |
| --- | --- | --- |
| Gender | 50 | Gender was self-reported at age 50 (man vs woman). |
| Social class at birth | 0 | Occupation of the father at the time of the participants’ birth was classified using the Registrar General’s Social Class Schema as ‘I professional’, ‘II managerial and technical’, ‘IIINM skilled nonmanual’, ‘IIIM skilled manual’, ‘IV semi-skilled manual’ or ‘V unskilled manual’); with those classified as “missing” who had unclassifiable occupation/had insufficient information/served in armed forces/were unemployed or sick or retired. The variable was binarised to indicate non-manual (classes I-IIINM) vs manual (classes IIIM-V) social class. |
| Conscientiousness | 16 | Following previously published approach (Atkins *et al.*, 2020), this measure was based on three facets of conscientiousness (hardworking, rigidity, cautiousness) from teacher-reported measures of attitudes recorded at age 16. Teachers rated students on three scales (ranging from 1 to 5): lazy-hardworking, flexible-rigid and impulsive-cautious. Summing up rating on all three facets resulted in the measure of conscientiousness ranging from 3 to 15, with the higher score indicating higher conscientiousness. |
| Cognitive ability | 11 | The cognitive ability was assessed using the General Ability Test (Douglas, 1964) at age 11, which comprised tests of both verbal and non-verbal skills. Scores from this test correlate strongly with IQ-type test scores (r=0.93), hence providing a good proxy for IQ scores (Douglas, 1964). Both verbal and non-verbal components were summed up, resulting with the total measure of cognitive ability ranging from 0 to 80, with the higher score indicating higher ability. |
| Internalising/  externalising problems | 16 | The subscales corresponding to internalising and externalising problems of the modified version of the Rutter A scale were used. The measure was completed by mothers of the participants as part of the home interview (Rutter *et al.*, 1970). Both externalising problems (e.g., “destroys own or others belongings”) and internalising problems (e.g., worries about many things) included four items. Each item had a 3-point response scale (“not true”=1, “somewhat true”=2, “certainly true”=3). Summing up items resulted in scores ranging from 4 to 12, with higher scores reflecting more externalising or internalising problems. The measure was tested in general population; acceptable inter-rater reliability (r = 0.64) and retest reliability (r = 0.74) (Rutter *et al.*, 1970). |
| Physical health problem | 16 | The variable captured physical health at age 16. It included information on currently having an abnormality in the main organ systems, such as cardiovascular or neurological. This was based on a full medical examination from a Local Authority Medical Officer, who completed a medical schedule and used information from medical records. |
| Parental involvement | 7 | Parental involvement was constructed in previously published research (Barboza Solís *et al.*, 2016). It comprised five items evaluating the time spend with the child (mum outing, dad outing, mum reading, dad reading and paternal role in the management of the child). These items were coded 2 if the activity occurred every week, 1 if occasionally and 0 if hardly ever. All items were summed up resulting in the score between 0 and 10, with the higher value indicating greater parental involvement. |

### eAppendix 1. Analytical approach to modelling average age trajectories of psychological distress.

We started with including age polynomials up to a cubic term and retaining them if their coefficients did not equal zero according to the Wald test (at p<0.05). This was due to previously found non-linearity of growth of psychological distress in adulthood.(Gondek *et al.*, 2021) Subsequently, the ACE score variable was added to the model, along with terms representing its interaction with age. This allowed the growth of distress to vary depending on the number of experienced ACEs. Subsequently, we added all covariates including gender, father’s occupational social class, maternal education, birthweight, gestational age, maternal age at birth and breastfeeding duration. We assessed if trajectories of distress differed across groups of ACEs with the Wald test (at p<0.05), by testing if all interaction coefficients combined – ACEs*age, ACEs*age^2^, ACEs*age^3^ – equalled zero. Finally, we included random linear age slope in the model allowing for heterogeneity in the trajectories of psychological distress. Models included non-linear age terms in the random part of the model did not converge. Hence, the fixed part of the final models included age (with non-linear terms), ACE variable, interaction terms for ACEs*age, covariates and the intercept (age 23). The random part of the final model captured variance in the intercept and age slope. A similar strategy was followed for prospectively reported ACEs, retrospectively reported ACEs, combined prospectively and retrospectively reported ACE scores and individual ACEs.

### eAppendix 2. Analytical approach to missing data.

At least one prospectively reported ACE was missing among n = 3,160 (36.0%) and at least one measure of the Malaise Inventory (the outcome) was missing in 2,853 (32.5%) individuals. The missing values were replaced with the multiple imputation. In line with recommendations we included all the variables from the analysis in the imputation model in order to preserve the relationship between the variables (Moons *et al.*, 2006, White *et al.*, 2011). To ensure that the model is compatible with the analysis, the interaction terms were included (Tilling *et al.*, 2016), treating them “just another variable” (the JAV approach) (Seaman S.R. *et al.*, 2012). However, as individual ACEs may contain more information than summative ACE scores, they were included in the imputation and after the imputation ACE scores were derived. Imputed ACE scores had, however, closely comparable prevalence.

The multiple imputation returns unbiased results under the missing at random (MAR) assumption (Collins *et al.*, 2001, Little and Rubin, 2002). The MAR mechanism, which is largely untestable, implies that systematic differences between the missing and the observed values can be explained by observed data (Collins *et al.*, 2001). The plausibility of the MAR assumption was maximised by including auxiliary variables, improving accuracy of the MI and minimising non-random variation in the imputed values (Sterne *et al.*, 2009). These variables included housing tenure at age 7, financial hardship at age 11, receiving free school meals at age 11. As shown by eTable 4, these variables were strongly associated with having missing data and with psychological distress. In addition, the imputation model was very rich due as psychological distress (the Malaise Inventory score) at preceding collection wave was the strongest predictor of having missing data at the subsequent collection waves (see eTable 4). Likewise, including other measures of mental health, such as the General Health Questionnaire-12 or the Clinical Interview Schedule – Revised, further strengthened the model.

The missing data were imputed using multiple imputation by chained equations (MICE), due to non-monotone pattern of missing values, and due to its ability to accommodate various types of variables in the imputation model, including continuous and categorical ones. This approach uses a series of univariate conditional imputation models to impute missing data (van Buuren, 2007). Continuous variables were imputed using predictive mean matching and categorical ones using logistic regressions. Predictive mean matching approach provides robust estimates if the normality assumption is in question (Morris *et al.*, 2014), as for instance with mental health outcomes that tend to be positively skewed (Counsell *et al.*, 2011), or when associations are non-linear (Morris *et al.*, 2014).

Laten class growth analysis was conducted using Full Information Maximum Likelihood (FIML) (Asparouhov and Muthen, 2014). FIML accounts for missing information, hence individuals with at least one measure of psychological distress were included. FIML assumes that data are under the missing at random assumption (MAR), hence auxiliary variables that predict missingness and the outcome (father’s occupational social class at birth, birthweight, mother’s age at birth and mother’s education) were included (see eTable 4) (Asparouhov and Muthen, 2014).

| eTable 4. Predictors of having missing data (vs not having missing data) and of psychological distress at age 50. | | |
| --- | --- | --- |
|  | Missing | Distress |
|  | RR (95%CI) | B (95%CI) |
| Housing tenure at age 7 (owner – reference category) |  | |
| Rented from council | 1.12 (1.06, 1.18) | 0.22 (0.12, 0.32) |
| Rented privately | 1.12 (1.04, 1.21) | 0.24 (0.09, 0.39) |
| Other | 1.05 (0.95, 1.17) | -0.15 (-0.35, 0.05) |
| Men (women – reference category) | 1.08 (1.04, 1.13) | -0.59 (-0.67, -0.50) |
| Non-manual father’s occupational class at age 0 (manual – reference) | 1.09 (1.04, 1.14) | 0.18 (0.09, 0.28) |
| Overcrowding at age 7 (no – reference category) | 1.13 (1.06, 1.21) | 0.28 (0.14, 0.42) |
| Psychological distress at age 23 | 1.16 (1.07, 1.26) | 2.06 (1.90, 2.23) |
| Financial hardship in previous year at age 11 (no – reference category) | 1.25 (1.16, 1.34) | 0.37 (0.20, 0.54) |
| Child receives free school meals at age 11 (no – reference category) | 1.25 (1.16, 1.34) | 0.42 (0.23, 0.60) |

| eTable 5. The estimates from covariates-adjusted multi-level model with the prospectively reported ACEs as the exposure. | | | |
| --- | --- | --- | --- |
|  | Without covariates |  | With covariates |
|  | B (95% CI) |  | B (95% CI) |
| Intercept | 1.058 (1.015 to 1.100) |  | 0.626 (-0.105 to 1.358) |
| Age | -0.126 (-0.139 to -0.112) |  | -0.126 (-0.139 to -0.113) |
| Age^2^ | 0.014 (0.012 to 0.015) |  | 0.014 (0.012 to 0.015) |
| Age^3^ | -0.000 (-0.000 to -0.000) |  | -0.000 (-0.000 to -0.000) |
| ACE score |  |  |  |
| 0 ACEs | Reference |  | Reference |
| 1 ACE | 0.308 (0.190 to 0.425) |  | 0.300 (0.190 to 0.409) |
| 2+ ACEs | 0.677 (0.498 to 0.856) |  | 0.652 (0.488 to 0.816) |
| *Interaction terms (ACE score*age)†* |  |  |  |
| 0 ACEs*Age | Reference |  | Reference |
| 1 ACE*Age | 0.037 (0.002 to 0.072) |  | 0.037 (0.004 to 0.070) |
| 2+ ACEs*Age | 0.026 (-0.029 to 0.081) |  | 0.026 (-0.026 to 0.078) |
| 0 ACEs*Age^2^ | Reference |  | Reference |
| 1 ACE*Age^2^ | -0.005 (-0.008 to -0.001) |  | -0.005 (-0.008 to -0.002) |
| 2+ ACEs*Age^2^ | -0.005 (-0.011 to 0.000) |  | -0.005 (-0.010 to 0.000) |
| 0 ACEs*Age3 | Reference |  | Reference |
| 1 ACE*Age^3^ | 0.000 (0.000 to 0.000) |  | 0.000 (0.000 to 0.000) |
| 2+ ACEs*Age^3^ | 0.000 (0.000 to 0.000) |  | 0.000 (0.000 to 0.000) |
| *Covariates* |  |  |  |
| Gender |  |  |  |
| Man |  |  | Reference |
| Woman |  |  | 0.649 (0.595 to 0.704) |
| Father's social class (at birth) |  |  |  |
| I professional |  |  | Reference |
| II managerial & technical |  |  | 0.026 (-0.113 to 0.164) |
| IIINM skilled non-manual |  |  | 0.058 (-0.091 to 0.208) |
| IIIM skilled manual |  |  | 0.120 (-0.010 to 0.251) |
| IV semi-skilled manual |  |  | 0.160 (0.011 to 0.308) |
| V unskilled manual |  |  | 0.276 (0.115 to 0.437) |
| Breastfeeding (reported at age 7) |  |  |  |
| No |  |  | Reference |
| Up to 1 month |  |  | -0.082 (-0.159 to -0.005) |
| Longer than 1 month |  |  | -0.085 (-0.153 to -0.017) |
| Mother's education (at birth) |  |  |  |
| Stayed beyond min leaving age |  |  | Reference |
| Left before min leaving age |  |  | 0.102 (0.036 to 0.169) |
| Birthweight (grams) |  |  | -0.000 (-0.000 to 0.000) |
| Maternal age at birth (years) |  |  | -0.003 (-0.007 to 0.002) |
| Gestational age (days) |  |  | 0.000 (-0.105 to 1.358) |
| *Random effects* |  |  |  |
| Intercept variance | 1.313 (1.259 to 1.369) |  | 0.893 (0.834 to 0.957) |
| Slope variance |  |  | 0.002 (0.001 to 0.002) |
| Within-individuals variance | 1.368 (1.340 to 1.397) |  | 1.162 (1.135 to 1.190) |
| † Global Wald test of interaction: without covariates, p = 0.001; with covariates, p = 0.0005. | | | |

| eTable 6. The estimates from covariates-adjusted multi-level model with the retrospectively reported ACEs as the exposure. | | | |
| --- | --- | --- | --- |
|  | Without covariates |  | With covariates |
|  | B (95% CI) |  | B (95% CI) |
| Intercept | 0.954 (0.907 to 1.000) |  | 0.383 ( -0.338 to 1.105) |
| Age | -0.115 (-0.130 to -0.100) |  | -0.115 (-0.129 to -0.101) |
| Age^2^ | 0.012 (0.011 to 0.013) |  | 0.012 (0.011 to 0.013) |
| Age^3^ | -0.000 (-0.000 to -0.000) |  | -0.000 (-0.000 to -0.000) |
| ACE score |  |  |  |
| 0 ACEs | Reference |  | Reference |
| 1 ACE | 0.293 (0.190 to 0.397) |  | 0.267 (0.174 to 0.359) |
| 2 ACEs | 0.456 (0.327 to 0.585) |  | 0.429 (0.314 to 0.545) |
| 3 ACEs | 0.696 (0.506 to 0.887) |  | 0.624 (0.452 to 0.797) |
| 4+ ACEs | 0.851 (0.706 to 0.996) |  | 0.755 (0.625 to 0.886) |
| *Interaction terms (ACE score*age)†* |  |  |  |
| 0 ACEs*Age | Reference |  | Reference |
| 1 ACE*Age | -0.012 (-0.045 to 0.021) |  | -0.012 (-0.043 to 0.019) |
| 2 ACEs*Age | -0.022 (-0.062 to 0.019) |  | -0.022 (-0.059 to 0.016) |
| 3 ACEs*Age | -0.019 (-0.077 to 0.040) |  | -0.019 (-0.074 to 0.036) |
| 4+ ACEs*Age | 0.019 (-0.030 to 0.068) |  | 0.019 (-0.027 to 0.065) |
| 0 ACEs*Age^2^ | Reference |  | Reference |
| 1 ACE*Age^2^ | 0.001 (-0.002 to 0.004) |  | 0.001 (-0.002 to 0.004) |
| 2 ACEs*Age^2^ | 0.002 (-0.002 to 0.006) |  | 0.002 (-0.001 to 0.006) |
| 3 ACEs*Age^2^ | 0.002 (-0.003 to 0.008) |  | 0.002 (-0.003 to 0.008) |
| 4+ ACEs*Age^2^ | -0.000 (-0.005 to 0.004) |  | -0.000 (-0.005 to 0.004) |
| 0 ACEs*Age3 | Reference |  | Reference |
| 1 ACE*Age^3^ | -0.000 (-0.000 to 0.000) |  | -0.000 (-0.000 to 0.000) |
| 2 ACEs*Age^3^ | -0.000 (-0.000 to 0.000) |  | -0.000 (-0.000 to 0.000) |
| 3 ACEs*Age^3^ | -0.000 (-0.000 to 0.000) |  | -0.000 (-0.000 to 0.000) |
| 4+ ACEs*Age^3^ | -0.000 (-0.000 to 0.000) |  | -0.000 (-0.000 to 0.000) |
| *Covariates* |  |  |  |
| Gender |  |  |  |
| Man |  |  | Reference |
| Woman |  |  | 0.600 (0.546 to 0.654) |
| Father's social class (at birth) |  |  |  |
| I professional |  |  | Reference |
| II managerial & technical |  |  | 0.043 (-0.092 to 0.179) |
| IIINM skilled non-manual |  |  | 0.075 (-0.071 to 0.221) |
| IIIM skilled manual |  |  | 0.126 (-0.001 to 0.253) |
| IV semi-skilled manual |  |  | 0.180 (0.036 to 0.325) |
| V unskilled manual |  |  | 0.285 (0.129 to 0.442) |
| Breastfeeding (reported at age 7) |  |  |  |
| No |  |  | Reference |
| Up to 1 month |  |  | -0.080 (-0.157 to -0.004) |
| Longer than 1 month |  |  | -0.087 (-0.154 to -0.021) |
| Mother's education (at birth) |  |  |  |
| Stayed beyond min leaving age |  |  | Reference |
| Left before min leaving age |  |  | 0.125 (0.060 to 0.190) |
| Birthweight (grams) |  |  | -0.000 (-0.000 to 0.000) |
| Maternal age at birth (years) |  |  | -0.000 (-0.005 to 0.005) |
| Gestational age (days) |  |  | 0.001 (-0.002 to 0.003) |
| *Random effects* |  |  |  |
| Intercept variance (between-individuals) | 1.240 (1.188 to 1.294) |  | 0.860 (0.802 to 0.922) |
| Slope variance (between-individuals) |  |  | 0.002 (0.001 to 0.002) |
| Within-individuals variance | 1.368 (1.340 to 1.397) |  | 1.164 (1.137 to 1.192) |
| † Global Wald test of interaction: without covariates, p = 0.06; with covariates, p = 0.12. | | | |

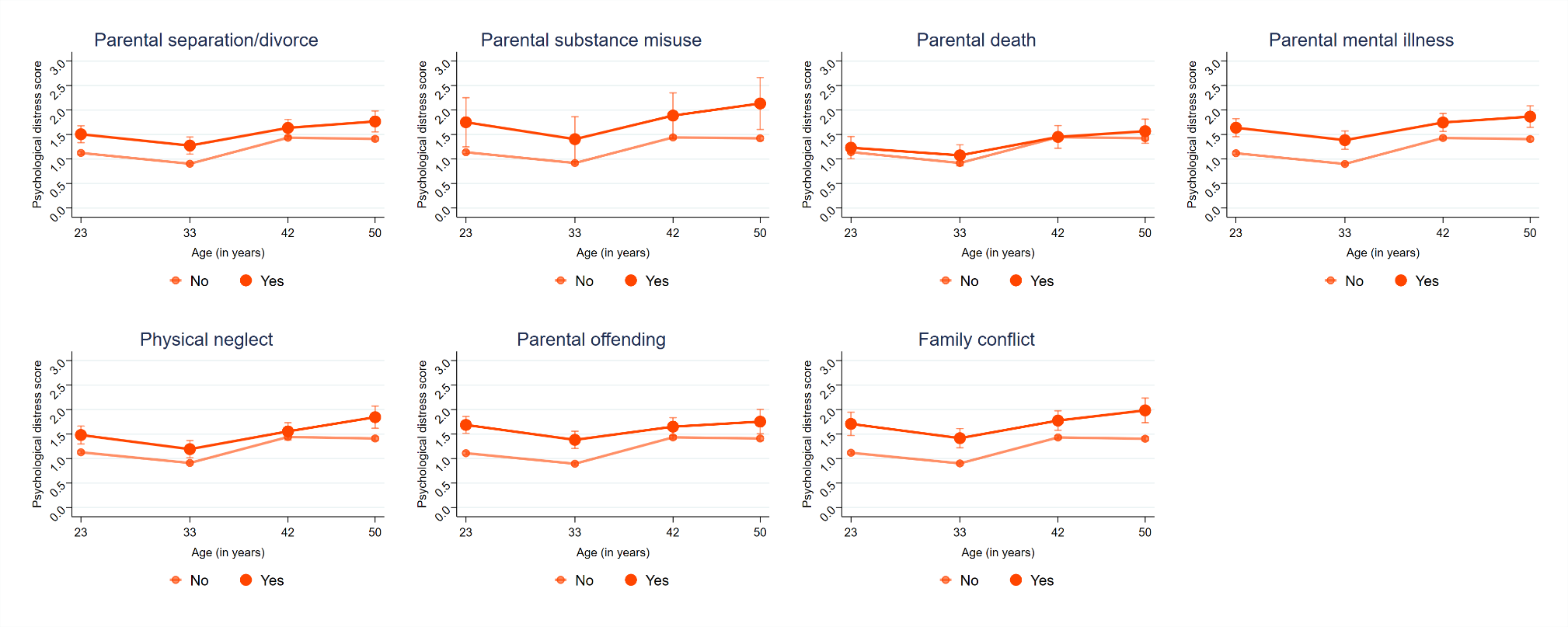

### eFigure 2. Trajectories of psychological distress across participants with individual prospectively reported ACEs.

*Note.* The estimates come from the model adjusted for covariates – gender, father’s occupational social class, maternal education, birthweight, gestational age, maternal age at birth and breastfeeding duration.

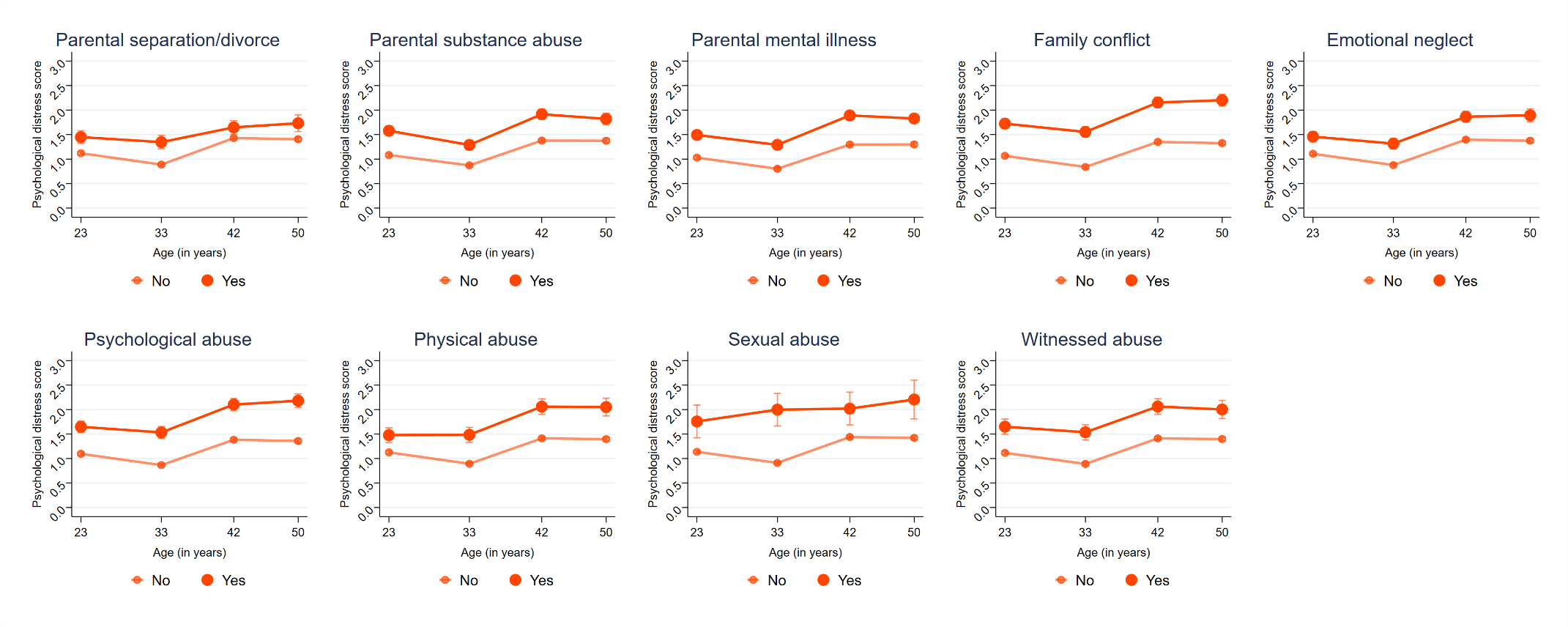

### eFigure 3. Trajectories of psychological distress across participants with individual retrospectively reported ACEs.

*Note.* The estimates come from the model adjusted for covariates – gender, father’s occupational social class, maternal education, birthweight, gestational age, maternal age at birth and breastfeeding duration.

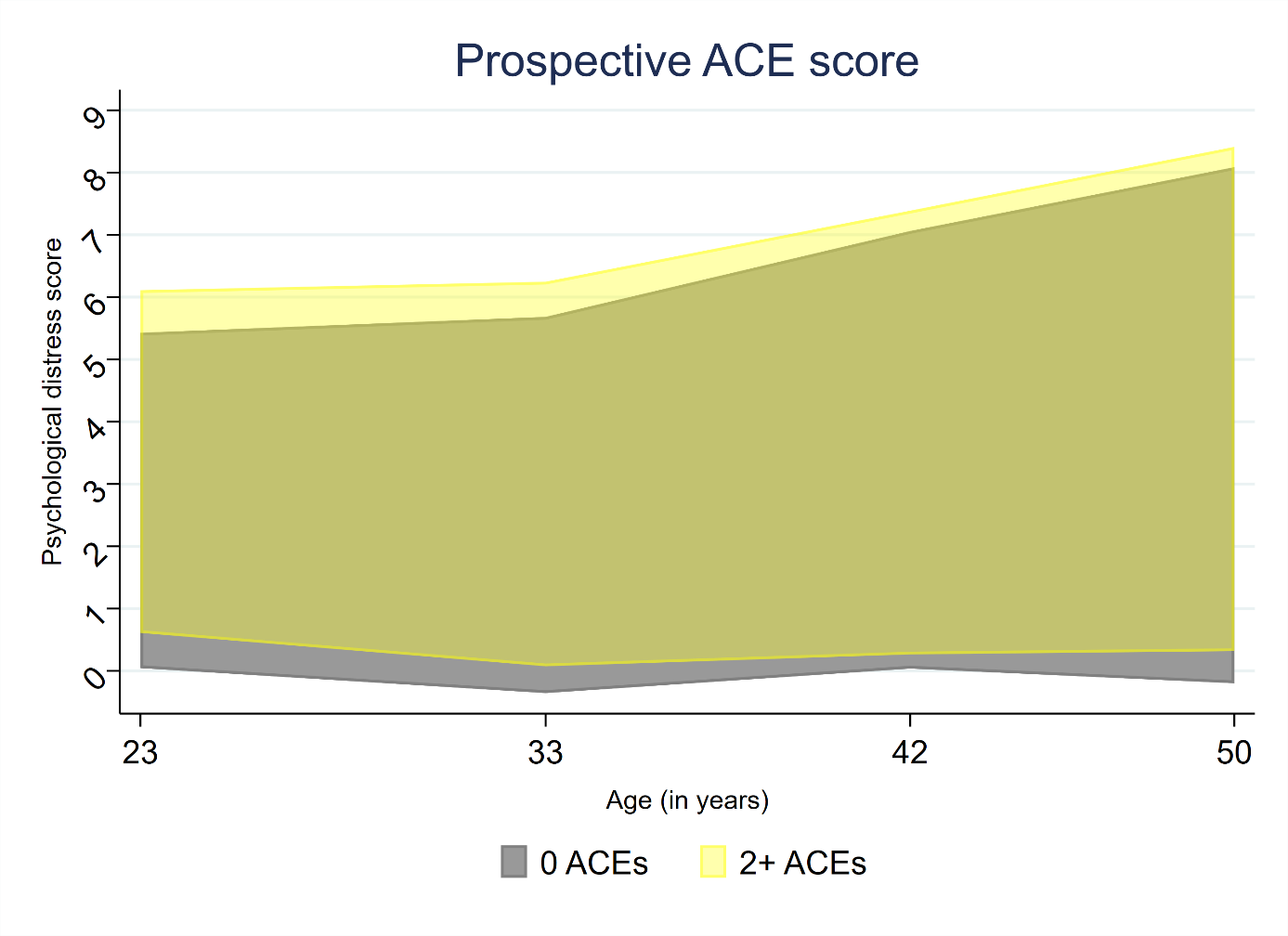

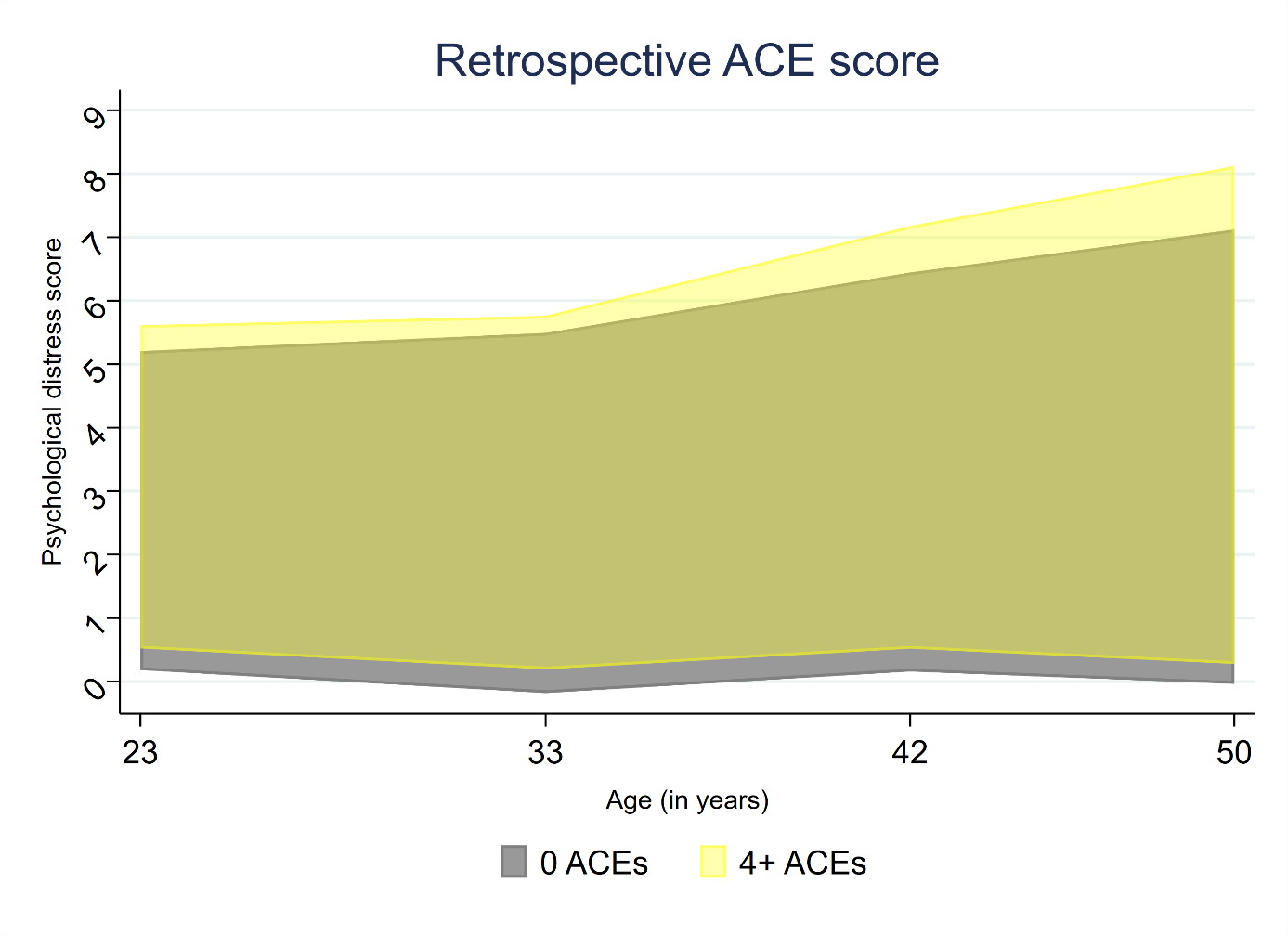

eFigure 4. Span of individual age trajectories of psychological distress.

*Note.* These show the extents to which individuals with and without ACEs vary in their trajectories of psychological distress (conditional on covariates).

| eTable 7. Comparison of models with different numbers of classes obtained with latent class growth analysis. | | | | |
| --- | --- | --- | --- | --- |
| **n = 8055** |  |  |  |  |
|  | **2 classes** | **3 classes** | **4 classes** | **5 classes** |
| AIC | 101461.427 | 99141.036 | 97484.093 | 96604.216 |
| BIC | 101538.361 | 99245.947 | 97616.980 | 96765.079 |
| Sample-adjusted BIC | 101503.405 | 99198.280 | 97556.602 | 96691.990 |
| Entropy | 0.912 | 0.881 | 0.883 | 0.882 |
| LMR-LRT | <0.001 | <0.001 | <0.001 | 0.0056 |
| VLMR-LRT | <0.001 | <0.001 | <0.001 | 0.0056 |
| % of the smallest class | 15.16% | 5.02% | 4.74% | 4.28% |
| AIC = Akaike Information Criterion; BIC = Bayesian Information Criterion; (V)LMR-LRT = (Vuong) Lo–Mendell–Rubin likelihood ratio tests. | | | | |

| eTable 8. Relative risk of belonging to four subgroups of age trajectories of psychological distress. | | | | |
| --- | --- | --- | --- | --- |
|  | Increasing symptoms in midlife (vs low symptoms) | | Moderate symptoms (vs low symptoms) | High symptoms  (vs low symptoms) |
|  | Relative risk ratio | | Relative risk ratio | Relative risk ratio |
| *Prospective ACE score* |  | |  |  |
| 0 ACEs (reference) | - | | - | - |
| 1 ACE | 1.08 (0.84 to 1.39) | | 1.34 (1.09 to 1.65) | 2.20 (1.63 to 2.96) |
| 2+ ACEs | 1.75 (1.27 to 2.41) | | 1.90 (1.42 to 2.54) | 3.31 (2.21 to 4.96) |
| *Retrospective ACE score* |  |  |  |  |
| 0 ACEs (reference) | - | | - | - |
| 1 ACE | 1.84 (1.50 to 2.27) | | 1.50 (1.24 to 1.81) | 2.05 (1.52 to 2.76) |
| 2 ACEs | 1.69 (1.30 to 2.19) | | 1.85 (1.49 to 2.31) | 2.35 (1.66 to 3.33) |
| 3 ACEs | 2.12 (1.50 to 3.02) | | 2.72 (2.05 to 3.60) | 4.63 (3.14 to 6.81) |
| 4+ ACEs | 2.56 (1.95 to 3.35) | | 2.79 (2.22 to 3.52) | 5.76 (4.24 to 7.82) |
| *Note.* The estimates are adjusted for covariates – gender, father’s occupational social class, maternal education, birthweight, gestational age, maternal age at birth and breastfeeding duration. | | | | |

| eTable 9. Relative risk of belonging to four subgroups of age trajectories of psychological distress using posterior probabilities from latent class growth analysis as regression weights. | | | | |
| --- | --- | --- | --- | --- |
|  | Increasing symptoms in midlife (vs low symptoms) | | Moderate symptoms (vs low symptoms) | High symptoms  (vs low symptoms) |
|  | Relative risk ratio | | Relative risk ratio | Relative risk ratio |
| *Prospective ACE score* |  | |  |  |
| 0 ACEs (reference) | - | | - | - |
| 1 ACE | 1.13 (0.91 to 1.39) | | 1.32 (1.11 to 1.56) | 2.14 (1.61 to 2.84) |
| 2+ ACEs | 1.75 (1.33 to 2.29) | | 1.82 (1.44 to 2.32) | 3.27 (2.23 to 4.80) |
| *Retrospective ACE score* |  |  |  |  |
| 0 ACEs (reference) | - | | - | - |
| 1 ACE | 1.74 (1.45 to 2.08) | | 1.44 (1.23 to 1.69) | 2.07 (1.56 to 2.75) |
| 2 ACEs | 1.73 (1.38 to 2.15) | | 1.81 (1.51 to 2.17) | 2.47 (1.77 to 3.43) |
| 3 ACEs | 2.06 (1.54 to 2.77) | | 2.62 (2.08 to 3.30) | 4.42 (3.04 to 6.43) |
| 4+ ACEs | 2.52 (2.00 to 3.18) | | 2.63 (2.16 to 3.21) | 5.73 (4.27 to 7.71) |
| *Note.* The estimates are adjusted for covariates – gender, father’s occupational social class, maternal education, birthweight, gestational age, maternal age at birth and breastfeeding duration. | | | | |

| eTable 10. Relative risk of belonging to four subgroups of age trajectories of psychological distress using less restrictive sample (n=13,130). | | | | |
| --- | --- | --- | --- | --- |
|  | Increasing symptoms in midlife (vs low symptoms) | | Moderate symptoms (vs low symptoms) | High symptoms  (vs low symptoms) |
|  | Relative risk ratio | | Relative risk ratio | Relative risk ratio |
| *Prospective ACE score* |  | |  |  |
| 0 ACEs (reference) | - | | - | - |
| 1 ACE | 1.16 (0.93 to 1.43) | | 1.34 (1.12 to 1.59) | 2.00 (1.62 to 2.46) |
| 2+ ACEs | 1.70 (1.29 to 2.24) | | 2.01 (1.64 to 2.45) | 3.86 (3.05 to 4.90) |
| *Retrospective ACE score* |  |  |  |  |
| 0 ACEs (reference) | - | | - | - |
| 1 ACE | 1.54 (1.26 to 1.87) | | 1.53 (1.29 to 1.82) | 1.99 (1.52 to 2.92) |
| 2 ACEs | 1.60 (1.26 to 2.04) | | 1.89 (1.54 to 2.33) | 2.76 (1.72 to 3.66) |
| 3 ACEs | 1.99 (1.45 to 2.73) | | 2.78 (2.13 to 3.63) | 4.66 (2.66 to 6.32) |
| 4+ ACEs | 2.43 (1.90 to 3.10) | | 3.31 (2.69 to 4.08) | 7.75 (5.87 to 10.23) |
| *Note.* The estimates are adjusted for covariates – gender, father’s occupational social class, maternal education, birthweight, gestational age, maternal age at birth and breastfeeding duration.  The sample included those who were not permanent emigrants, were alive by age 50 and had at least two measures of prospectively reported adverse childhood experiences (n=13,130). | | | | |

| eTable 11. Relative risk of belonging to four subgroups of age trajectories of psychological distress. | | |
| --- | --- | --- |
|  | Increasing symptoms in midlife (vs low symptoms) | Moderate/high symptoms (vs low symptoms) |
| *Effect modifiers as exposures* | Relative risk ratio | Relative risk ratio |
| Gender | - | - |
| Woman (vs man) | 2.18 (1.86 to 2.56) | 2.60 (2.30 to 2.94) |
| Social class at birth* | - | - |
| Manual (vs non-manul) | 1.15 (0.96 to 1.37) | 1.27 (1.11 to 1.46) |
| Parental involvement at age 7** | 0.93 (0.89 to 0.96) | 0.93 (0.90 to 0.95) |
| Conscientiousness at age 11** | 0.97 (0.92 to 1.02) | 0.93 (0.90 to 0.96) |
| Cognitive ability at age 11** | 0.97 (0.96 to 0.99) | 0.96 (0.95 to 0.97) |
| Internalising problems at age 16** | 1.15 (1.09 to 1.21) | 1.22 (1.17 to 1.26) |
| Externalising problems at age 16** | 1.14 (1.05 to 1.23) | 1.32 (1.25 to 1.40) |
| Physical health problem at age 16** | - | - |
| Yes (vs no) | 1.34 (1.07 to 1.77) | 1.30 (1.10 to 1.55) |
| *The estimates are adjusted for gender.  **The estimates are adjusted for gender, father’s occupational social class at birth, maternal education, birthweight, gestational age, maternal age at birth and breastfeeding duration. | | |

**References**

**Asparouhov, T. & Muthen, B.** (2014). Auxiliary Variables in Mixture Modeling: Three-Step Approaches Using Mplus. *Structural Equation Modeling* **21**, 329-41.

**Atkins, R., Turner, A. J., Chandola, T. & Sutton, M.** (2020). Going beyond the mean in examining relationships of adolescent non-cognitive skills with health-related quality of life and biomarkers in later-life. *Economics & Human Biology* **39**, 100923.

**Barboza Solís, C., Fantin, R., Castagné, R., Lang, T., Delpierre, C. & Kelly-Irving, M.** (2016). Mediating pathways between parental socio-economic position and allostatic load in mid-life: Findings from the 1958 British birth cohort. *Social Science & Medicine* **165**, 19-27.

**Collins, L. M., Schafer, J. L. & Kam, C. M.** (2001). A comparison of inclusive and restrictive strategies in modern missing data procedures. *Psychological Methods* **6**, 330-351.

**Counsell, N., Cortina-Borja, M., Lehtonen, A. & Stein, A.** (2011). Modelling psychiatric measures using Skew-Normal distributions. *European Psychiatry* **26**, 112-114.

**Douglas, J. W. B.** (1964). *The Home and the School.* . McGibbon and Kee: London.

**Gondek, D., Bann, D., Patalay, P., Goodman, A., Richards, M. & Ploubidis, G. B.** (2021). Psychological distress from adolescence to early old age: Evidence from the 1946, 1958 and 1970 British birth cohorts. *Psychological Medicine*.

**Little, R. & Rubin, D. B.** (2002). *Statistical analysis with missing data*. Wiley: Hoboken, N.J.

**Moons, K. G. M., Donders, R. A. R. T., Stijnen, T. & Harrell, F. E. J.** (2006). Using the outcome for imputation of missing predictor values was preferred. *Journal of Clinical Epidemiology* **59**, 1092–101.

**Morris, T. P., White, I. R. & Royston, P.** (2014). Tuning multiple imputation by predictive mean matching and local residual draws. *BMC Med Res Methodol* **14**, 75.

**Rutter, M., Tizard, J. & Whitmore, K.** (1970). *Education, Health and Behaviour*. Longmans: London.

**Seaman S.R., Bartlett J.W. & I.R., W.** (2012). Multiple imputation of missing covariates with non-linear effects and interactions: an evaluation of statistical methods. *BMC Medical Research Methodology* **12**.

**Sterne, J. A., White, I. R., Carlin, J. B., Spratt, M., Royston, P., Kenward, M. G., Wood, A. M. & Carpenter, J. R.** (2009). Multiple imputation for missing data in epidemiological and clinical research: potential and pitfalls. *BMJ* **338**, b2393.

**Tilling, K., Williamson, E. J., Spratt, M., Sterne, J. A. & Carpenter, J. R.** (2016). Appropriate inclusion of interactions was needed to avoid bias in multiple imputation. *Journal of Clinical Epidemiology* **80**, 107–115.

**van Buuren, S.** (2007). Multiple imputation of discrete and continuous data by fully conditional specification. *Stat Methods Med Res.* **16**, 219–42.

**White, I. R., Royston, P. & Wood, A. M.** (2011). Multiple imputation using chained equations: issues and guidance for practice. *Statistics in Medicine* **30**, 377–99.
